# Population-based Risk of Psychiatric Disorders Associated with Recurrent CNVs

**DOI:** 10.1101/2023.09.04.23294975

**Authors:** Morteza Vaez, Simone Montalbano, Xabier Calle Sánchez, Kajsa-Lotta Georgii Hellberg, Saeid Rasekhi Dehkordi, Morten Dybdahl Krebs, Joeri Meijsen, John Shorter, Jonas Byberg-Grauholm, Preben B Mortensen, Anders D Børglum, David M Hougaard, Merete Nordentoft, Daniel H Geschwind, Alfonso Buil, Andrew J Schork, iPSYCH Investigators, Dorte Helenius, Armin Raznahan, Wesley K Thompson, Thomas Werge, Andrés Ingason

**Affiliations:** Institute of Biological Psychiatry, Mental Health Services, Copenhagen University Hospital, DK-4000 Roskilde, Denmark; The Lundbeck Foundation Initiative for Integrative Psychiatric Research (iPSYCH), Copenhagen and Aarhus, Denmark; Department of Science and Environment, Roskilde University, DK-4000 Roskilde, Denmark; Center for Neonatal Screening, Department for Congenital Disorders, Statens Serum Institut, DK-2300 Copenhagen, Denmark; National Centre for Register-based Research, Aarhus University, DK-8210 Aarhus, Denmark; Department of Biomedicine—Human Genetics and the iSEQ Center, Aarhus University, DK-8000 Aarhus, Denmark; Center for Genomics and Personalized Medicine, DK-8000 Aarhus, Denmark; Mental Health Center Copenhagen, Copenhagen University Hospital, DK-2400 Copenhagen, Denmark; Department of Neurology, University of California, Los Angeles, CA, USA; Department of Human Genetics, University of California, Los Angeles, CA, USA; Center for Autism Research and Treatment, Semel Institute, David Geffen School of Medicine at UCLA, University of California, Los Angeles, CA, USA; Program in Neurobehavioral Genetics, Semel Institute, David Geffen School of Medicine at UCLA, University of California, Los Angeles, CA, USA; Center for Human Development, University of California, San Diego, CA, USA; Lundbeck Foundation Center for GeoGenetics, GLOBE Institute, University of Copenhagen, DK-1350 Copenhagen, Denmark; Neurogenomics Division, The Translational Genomics Research Institute (TGEN), Phoenix, AZ, USA; See list of iPSYCH investigators and their affiliations in Supplementary Note 1; Section on Developmental Neurogenomics, Human Genetics Branch, National Institute of Mental Health Intramural Research Program, Bethesda, MD, USA; Laureate Institute for Brain Research, Tulsa, OK, USA; Department of Clinical Medicine, University of Copenhagen, DK-2200 Copenhagen, Denmark

## Abstract

Recurrent copy number variants (rCNVs) are associated with increased risk of neuropsychiatric disorders but their pathogenic population-level impact is unknown. We provide population-based estimates of rCNV-associated risk of neuropsychiatric disorders for 34 rCNVs in the iPSYCH2015 case-cohort sample (n=120,247).

Most observed significant increases in rCNV-associated risk for ADHD, autism or schizophrenia were moderate (HR:1.42-5.00), and risk estimates were highly correlated across these disorders, the most notable exception being high autism-associated risk with Prader-Willi/ Angelman Syndrome duplications (HR=20.8). No rCNV was associated with significant increase in depression risk. Also, rCNV-associated risk was positively correlated with locus size and gene constraint. Comparison with published rCNV studies suggests that prevalence of some rCNVs is higher, and risk of psychiatric disorders lower, than previously estimated.

In an era where genetics is increasingly being clinically applied, our results highlight the importance of population-based risk estimates for genetics-based predictions.

## Introduction

Recurrent copy number variants (rCNVs) arise through non-allelic homologous recombination mediated by low-copy repeat sequences. rCNVs are particularly relevant for human disease genetics studies because they are maintained at fairly stable population frequencies (due to high *de novo* mutation rates) despite often being associated with substantial pathogenicity^1,2^ and reduced reproductive fitness^3^. At least 40 loci in the human genome harbour rCNVs conferring moderate to high risk of intellectual disability (ID), developmental delay (DD) and congenital malformations (CM)^4^. Many of these have also been associated with increased risk of neuropsychiatric disorders; autism spectrum disorder (ASD)^1^, attention-deficit/hyperactivity disorder (ADHD)^5^, schizophrenia spectrum disorder (SSD)^6-8^ and epilepsy^9^, while evidence for associated risk of bipolar disorder (BPD)^10^ and major depressive disorder (MDD)^11^ is limited.

Most rCNV studies have derived estimates of prevalence and associated risk of disease from clinical case-control studies, often based on highly selected cases (e.g., with severe or long-term illness) and controls (e.g., screened for lack of any family history of mental illness). While this study design can increase power for detecting associations between genetic exposures and disease outcomes, it may yield distorted estimates of the prevalence of the exposure and the associated risk of the outcome. Alternative study designs that rely on community-based samples – most prominently the UK Biobank (UKB)^11-12^ – benefit from larger sample sizes and a multitude of available study outcomes. However, the UKB suffers a “healthy volunteer” bias^13^ that has been shown to affect association estimates in genetic studies^14,15^. The lack of population-unbiased estimates of rCNV prevalence and disease risk compromises proper implementation of genetically informed clinical care, including guiding patient assessment and choice of treatment, and genetic counselling.

Our recent research in Danish registers and biobanks^16^ found that deletions at 22q11.2 may have much more modest effects on risk of SSD in the Danish population^17,18^ than reported in clinical case-control studies^6,19-21^. This dampening of effect size estimates compared to case-control studies was also observed for SSD, ADHD and ASD when we applied the same population-based analyses to rCNVs at six other loci (1q21.1, 15q11.2, 15q13.3, 16p11.2, 17p12, and 17q12).^22^

In the present study, we expand on our initial population-based analyses of rCNV prevalence and penetrance in two key directions. First, we expand these analyses to systematically consider deletions and duplications across 30 rCNV loci most commonly associated with genomic disorders and clinical phenotypes.^12^ Second, we also compare the effects on risk across rCNV loci and type, and across psychiatric outcomes. Such direct comparisons are essential for clarifying if rCNVs impart risk profiles differently – an open question of central mechanistic and translational importance in psychiatry. We pursue both of these advances in a substantially increased sample size^23^ – affording unprecedented precision and power to characterise population-unbiased rCNV prevelances and penetrances.

## Results

### Quality control (QC) and analysis of CNV detection accuracy

We processed PennCNV^24^ calls for available samples through the QCtreeCNV pipeline^25^ and additional QC tests (Methods, Supplementary Note 2), yielding 3,589 verified rCNV calls in 120,247 samples; 79,535 cases (anyone having received a hospital diagnosis of any of the index psychiatric disorders, see Methods) and 43,311 from the subcohort (a randomly drawn sample from the same birth cohort, including an overlap [cases-in-subcohort] of 2,599 samples, see Methods). The sample genotype success rate corresponded to 85.8% of the 140,087 samples ascertained in the iPSYCH2015 design^23^ (Extended Data Figure 1), and did not differ across the case, subcohort, and cases-in-subcohort groups (Fisher’s Exact Test; simulated P=0.12). The sample included 64,735 (53.8%) males and 55,512 (46.2%) females, as recorded at birth. Age at end of follow-up (AEF) ranged between 7.0-34.7 years (mean 21.8, standard deviation 7.0). All participants were born in Denmark but information on self-declared ethnicity was not available. Out of 30 queried loci, 12 were excluded due to poor quality of SNP array data, low carrier count or failure to meet the proportionality of hazards assumption underlying the fitted Cox Proportional Hazards (CPH) models used to estimate rCNV-associated risk of iPSYCH2015 disorders (Methods, Supplementary Table 1).

### rCNV prevalence and comparison with UKB

We estimated population prevalence and 95% confidence intervals (CI95%) for each of 18 deletions and duplications in the full case-cohort sample using finite population correction (fpc) to account for oversampling of cases (Methods). Consistent with previous rCNV studies^12,26,27^ the overall prevalence of duplications was higher than that of deletions (1.29% vs 0.99%, P=2.2×10^−5^). However, the prevalence disparity differed substantially across loci (Figure 1A); five loci (TAR, 1q21.1, 15q13.3, 16p13.11, 16p11.2 and 22q11.2) had significantly higher prevalence of duplications and two loci (2q13 and 16p12.1) had significantly higher prevalence of deletions (Supplementary Table 2). We then compared rCNV prevalence between our study and the UKB^12^ for the 17 loci with estimates in both studies (Methods). The overall population-based prevalence of deletions was higher in iPSYCH2015 (0.98% vs 0.70%; P=7.7×10^−12^) whereas overall duplication prevalence was similar in both studies (1.27% vs 1.23%, P=0.42). In the per-locus comparison nine deletions and one duplication (16p11.2-dup) had higher prevalence (P_FDR_<0.05) in iPSYCH2015, while 13q12.12-dup was more prevalent in UKB, and 23 rCNVs showed no significant differences (Figure 1B, Supplementary Table 2).

**Figure 1:**
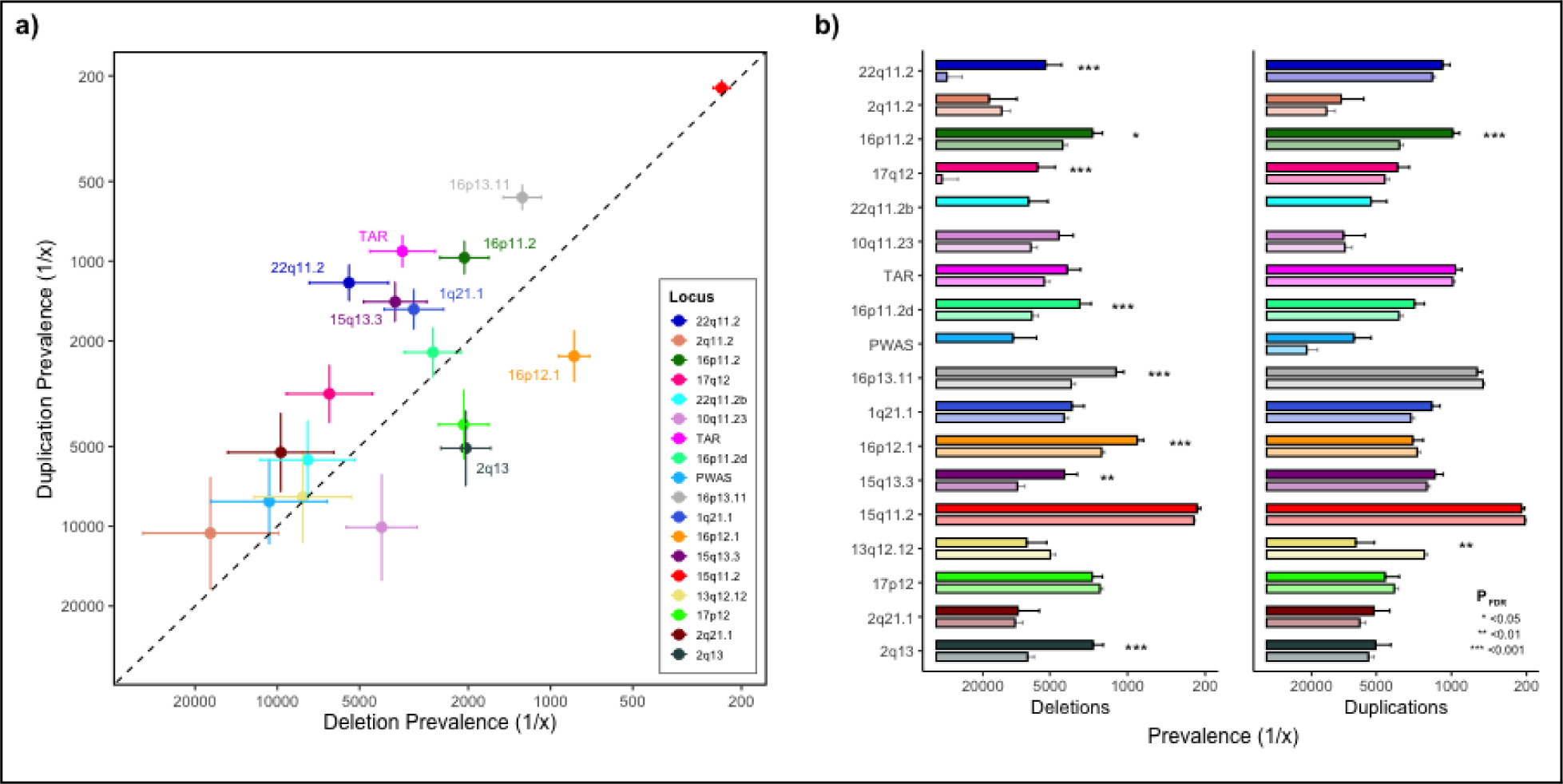
Population-based rCNV prevalence in iPSYCH2015 and comparison with UKB. **a)** Population-based prevalence in iPSYCH2015 of deletions (x-axis) and duplications (y-axis) at 18 rCNV loci. Dashed line indicates del-dup prevalence parity. The 8 loci with significant del-dup prevalence disparity (P_FDR_<0.05) are indicated on the plot. **b)** Population-based rCNV prevalence of deletions (left) and duplications (right) in iPSYCH2015 (full colour, above) compared to that reported for the UKB^12^ (semi-transparent colour, below), with loci ordered from top to bottom by decreasing sum LOEUF score^28^. In the UKB study, 22q11.2b (turquoise) was not included as an independent locus and the prevalence of PWAS deletion was <1/100,000^12^. Significant differences in prevalence between the two studies are indicated with asterisks (P_FDR_<0.05 (*), <0.01 (**), and <0.001 (***), respectively). Error bars indicate the standard error (SE) of the weighted population-based prevalence in iPSYCH2015 and observed prevalence in UKB^12^, respectively. For detailed results see Supplementary Table 1.

### CNV-associated risk of psychiatric disorders

We estimated population-based hazard ratios (HR) for each deletion and duplication at the 18 rCNV loci for each of the ascertained psychiatric diagnosis groups (Methods, Extended Data Figure 1). Per-diagnosis results for the main diagnosis groups (ASD, ADHD, MDD and SSD) are shown in Figure 2, while a per-locus version of all estimates, including for schizophrenia (ICD10: F20), BPD (for which HR could be estimated for only 11 rCNVs) and the broader diagnosis groups (any affective disorder and any iPSYCH2015 disorder, Methods) are provided in Extended Data Figure 2 and Supplementary Table 3.

**Figure 2:**
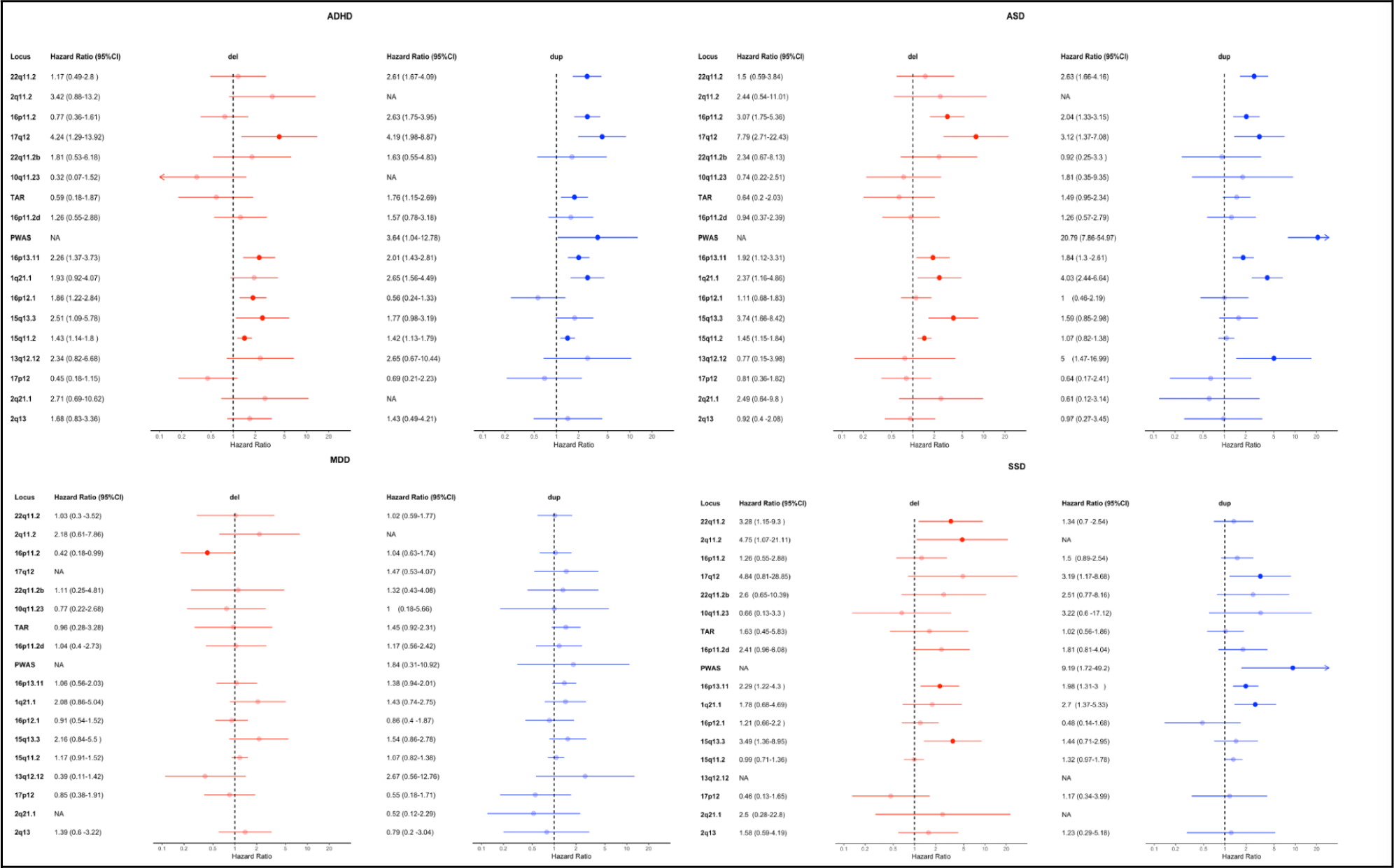
rCNV-associated risk of psychiatric disorders in the iPSYCH2105 case-cohort. rCNV-associated Hazard ratios (HR) and 95% confidence intervals (CI95%) were derived from Cox proportional hazards (CPH) models using inverse probability of sampling (IPS) weights, and are indicated for deletions and duplications by red and blue colour, respectively, for the 18 rCNV loci assessed in this study. Associated HRs with P<0.05 (i.e. with CI95% not overlapping HR=1) are bolded, and loci are ordered on the y-axis from top to bottom by decreasing sum LOEUF score^28^. Abbreviations for the four most common psychiatric disorders targeted by the iPSYCH2015 case-cohort design (depicted on plot) are as follows: ADHD; attention-deficit hyperactivity disorder, ASD; autism spectrum disorder, MDD; major depressive disorder, SSD; schizophrenia spectrum disorder. Full per-locus results, including estimates also for Schizophrenia (SCZ), bipolar disorder (BPD), any affective disorder (AFF), any iPSYCH disorder (ANY), intellectual disability (ID) and epilepsy, are provided in Extended Data Figure 2.

Risk estimates exceeded 1.5 for a majority of rCNVs tested for ASD (18 of 34), ADHD (21 of 32), and SSD (19 of 31), but only for 6 of 32 tested for MDD (Figure 2). In fact, we observed no significantly increased HRs for MDD (Figure 2) or the broader category of any affective disorder (Extended Data Figure 2). Six rCNVs (1q21.1-dup, Prader-Willi/Angelman Syndrome (PWAS) -dup, 15q13.3-del, 16p13.11-del, 16p13.11-dup and 17q12-dup) were associated with significantly increased HR of all three; ADHD, ASD, and SSD, and further four rCNVs (15q11.2-del, 16p11.2-dup, 17q12-del and 22q11.2-dup) with increased HR of both ADHD and ASD (Extended Data Figure 2). The greatest increases in HR were observed for PWAS-dup with ASD (HR=20.7; CI95%: 7.8-54.9) and SSD (HR=9.1; CI95%: 1.7-49.2), and for 17q12-del with ADHD (HR=4.2; CI95%: 1.2-13.9) (Figure 2).

We compared the association results for the four main diagnosis groups with the largest available published case-control studies. The number of rCNVs with available estimates in those studies ranged from 11 for ASD^1^ to 13 for ADHD^5^, 20 for MDD^11^, and 22 for SSD^6-8^. Risk estimates (HRs from our study and odds ratios (ORs) from comparison studies) were compared with a Welch’s t-test and adjusted for multiple comparisons for each disorder separately using false discovery rate (FDR). Risk estimates were significantly lower in our study for 1q21.1-del, 15q11.2-del, 16p11.2-dup, 17p12-del and 22q11.2-del for SSD, 16p11.2-del and 16p11.2-dup for ASD, and 22q11.2-del for ADHD (Extended Data Figure 3, Supplementary Table 4). The only significantly higher risk estimate in iPSYCH2015 was for 22q11.2-dup in SSD, for which we find no indication (HR=1.34; CI95%: 0.70-2.54) of the protective effect reported in the comparison study (OR=0.15; CI95%: 0.04-0.52)^6^. While HRs and ORs are slightly different estimates of risk, a sensitivity analysis found very high correlation across all diagnosis groups between the CPH-derived HRs and corresponding ORs derived from logistic regression models of iPSYCH2015 data (Pearson’s correlation test: r=0.94, P<2.2×10^−16^, Supplementary Table 5) indicating that they are comparable.

As no individual rCNV was significantly associated with increased risk of MDD, we assessed the overall effect of rCNV carrier status on MDD diagnosis using nested generalised linear models (GLMs) with and without a categorical rCNV variable (with separate levels for each rCNV and “no rCNV” as reference) and sex and AEF as covariates (Methods). A likelihood ratio test (LRT) found no effect of rCNV carrier status on MDD prediction (*χ*2=40.32; P=0.28); a stark contrast to corresponding results for ADHD, ASD, and SSD (*χ*2>104.68 and P<1.27×10^−8^ for each). Therefore, we excluded MDD from the following comparative analyses of rCNV-associated risk across diagnosis groups, between deletions and duplications, and across loci.

### Comparison of rCNV-associated risk across disorders

The availability of information on multiple outcomes and rCNVs carrier status in the same population-based dataset allowed us to test if rCNV-associated risk varied across outcomes, which we did through several complementary approaches. First, we plotted CNV-associated HR estimates for each pair of the three most relevant diagnosis groups (ADHD, ASD and SSD) and observed high pairwise correlations (r_(ASD-ADHD)_=0.68; P=2×10^−5^, r_(ASD-SSD)_=0.71; P=7×10^−6^,r_(ADHD-SSD)_=0.75; P=2×10^−6^; Figure 3A).

**Figure 3:**
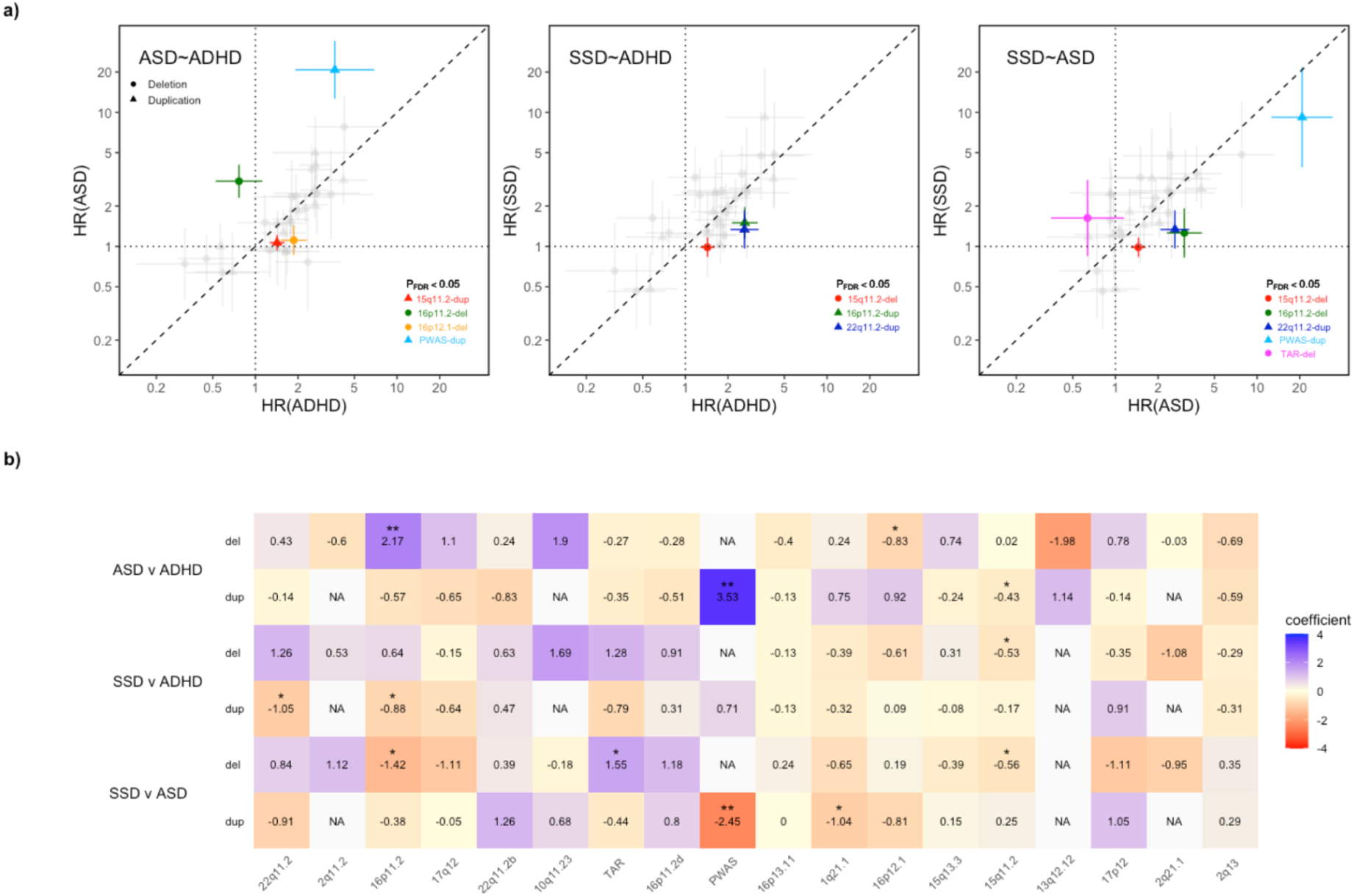
Contrasts in associated risk between main psychiatric outcomes across rCNVs. **a)** Hazard ratios (HR) with error bars indicating standard errors (SE) for rCNV deletions (circle) and duplications (triangle) are plotted in a pairwise comparison for ADHD-v-ASD (left), SSD-v-ASD (middle), and SSD-v-ADHD (right), with estimates for each disorder on the x- and y-axis, respectively. The dashed line indicates risk parity. On each pairwise comparison plot, we have highlighted those rCNVs showing significant evidence of a rCNV-by-diagnosis interaction in generalized estimating equations (GEE) predicting case status for two diagnoses at a time (P_FDR_<0.05), with colours corresponding to the rCNV locus in Figure 1. **b)** We constructed a heatmap of the coefficients from the rCNV-by-diagnosis interaction derived from GEE models for each pairwise comparison across all rCNVs. Significant coefficients are indicated with asterisks (P_FDR_<0.05 (*), P_FDR_<0.01 (**)). rCNV loci are ordered on the x-axis from left to right by decreasing sum LOEUF score^28^, with coefficients for deletions and duplications shown in separate rows for each pairwise diagnosis comparison. Only rCNVs with available HRs for both compared diagnoses were included in each GEE analysis.

To test for an omnibus rCNV-associated difference between pairs of disorders, we used generalised estimating equations (GEE) which allow accounting for individuals in multiple diagnosis groups, and fitted nested models with and without an interaction term between diagnosis group and overall rCNV carrier status (with sex at birth and AEF as covariates), with diagnosis of either of the two disorders as outcome (Methods). This revealed a significant difference in overall rCNV-associated effects between ASD and SSD, and ADHD and SSD, but not between ASD and ADHD (ASD vs SSD: *χ*^2^=7.47, P=0.0063; ADHD vs SSD: *χ*^2^=10.08, P=0.0015; ASD vs ADHD: *χ*^2^=0.21, P=0.64).

Then, we tested for rCNV-specific differences using the same approach but replacing the overall rCNV carrier variable with the categorical rCNV variable described above (previous subsection). This analysis showed significant effect of adding the rCNV-diagnosis interaction term in all three comparisons, suggesting that for each pair of disorders at least some rCNVs differ in effect size (*X*^2^ =94.6, P=4.2×10^−8^; *X*^2^ =72.9, P=3.1×10^−5^; *X*^2^ =50.1, P=1.2×10^−2^). Post hoc analyses of the interaction coefficients from the GEE analyses revealed no clear trend for rCNVs consistently conferring higher or lower risk of any of the three diagnoses compared with either of the other two (Figure 3B, Supplementary Table 6), but among rCNVs with significant interaction coefficients (P_FDR_<0.05) both PWAS-dup and 16p11.2-del were associated with higher HR for ASD than for ADHD or SSD, and both 15q11.2-del and 22q11.2-dup were associated with higher HR for ASD and ADHD than for SSD (Figure 3A).

### Comparison of risk between deletions and duplications

The distribution of HR estimates for deletions vs duplications across loci for the three main diagnosis groups reveals no indication of a consistent dosage-dependent effect (Figure 4). To formally test whether the level of rCNV-associated risk is dependent on dosage type (deletion or duplication), we used GEE models to predict case status from dosage type among rCNV carriers using AEF, sex and diagnosis group as covariates, for all 16 loci with risk estimates available for both deletions and duplications (Methods). This analysis found no omnibus effect of rCNV type across all three diagnosis groups (χ2=0.11, P=0.73). Also, a comparison of the GEE models with and without an interaction term between rCNV type and locus found no indication of significant locus-specific rCNV type effects (χ2=20.66, P=0.14).

**Figure 4:**
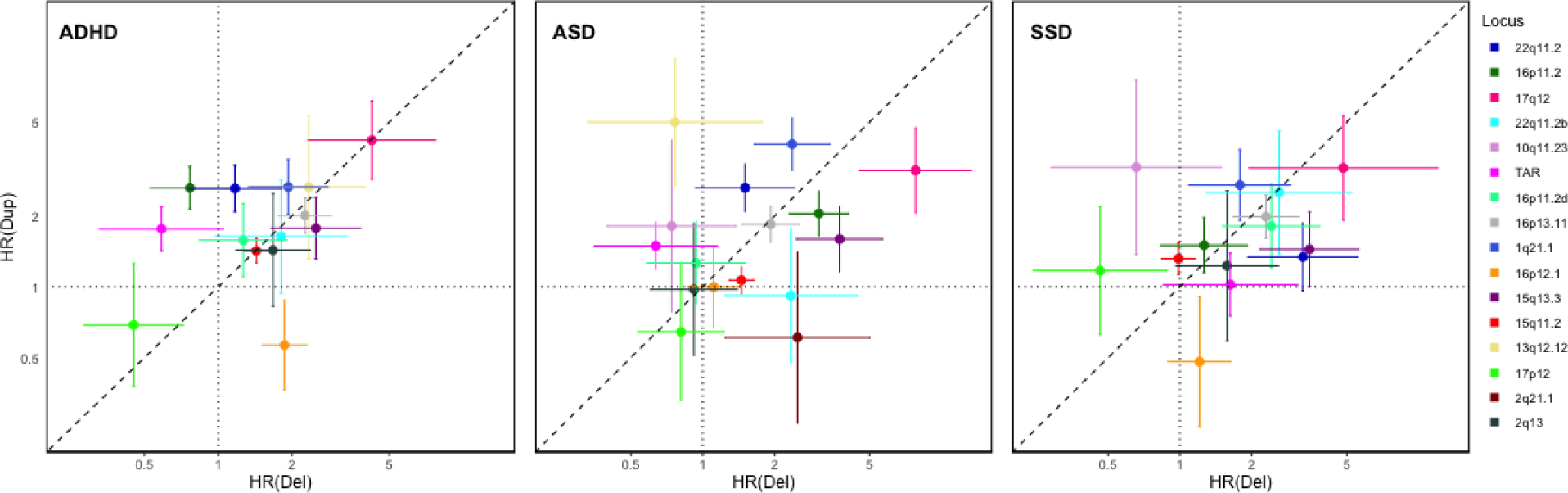
Contrasts in associated risk by dosage change across rCNV loci. Hazard ratios (HR) with error bars indicating standard errors (SE) for deletions and duplications are plotted on the x-axis and y-axis, respectively for ADHD, ASD and SSD. The 16 loci with available HR estimates for both deletions and duplications for at least one diagnosis are indicated by different colours (see figure legend) corresponding to previous figures. The dashed line indicates risk parity. We found no evidence of a locus-by-dosage interaction increasing prediction of case status in an omnibus test across all loci and diagnoses between a GEE model with the interaction term and a nested model without the interaction term (likelihood ratio test; P=0.16).

### Locus features as predictors of rCNV-associated risk

As HRs for ADHD, ASD and SSD vary significantly across rCNVs, we hypothesised that rCNV-associated risk would be correlated with the gene content or locus size, as potential indicators of pathogenicity. Also, given the significant differences in prevalence of some rCNVs between our study and the UKB^11^, we hypothesised that rCNV-associated risk would be correlated with their population-based prevalence and/or the difference in prevalence between our population-based study and the volunteer-based UKB study.

We plotted the HR estimates for the three disorders against locus size and locus-wide loss-of-function observed/expected upper bound fraction (LOEUF) score^28^ across loci (Figure 5 a-b), and population-based prevalence and the iPSYCH2015/UKB prevalence ratio across rCNVs (Figure 5 c-d), fitting log-linear trend lines for deletion- and duplication-associated estimates separately (excluding PWAS due to outlying locus size and HR estimates). While differing somewhat across diagnoses, the plots indicate that HRs tend to increase with locus size (Figure 5a), LOEUF score (Figure 5b) and iPSYCH/UKB rCNV prevalence ratio (Figure 5d), while decreasing with rCNV prevalence itself (Figure 5c).

**Figure 5:**
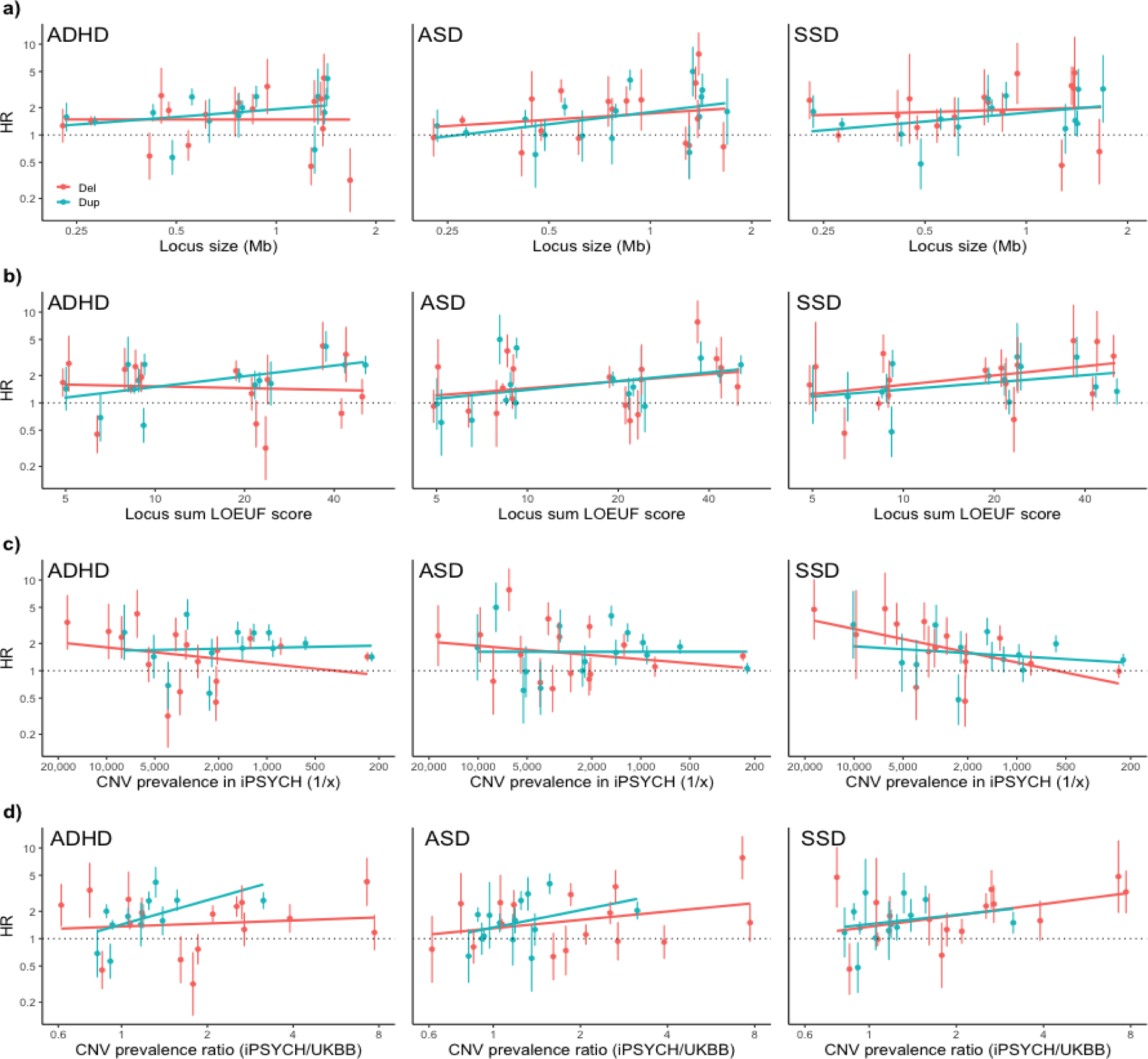
rCNV-associated risk as a function of locus properties and rCNV prevalence. Hazard ratios (HR) with error bars indicating standard errors (SE) for deletions (light red) and duplications (iris blue) associated with ADHD (left), ASD (middle), and SSD (right) are plotted against **a)** locus size and **b)** sum LOEUF score^28^, as well as **c)** population prevalence in iPSYCH2015 and **d)** the population prevalence ratio between iPSYCH2015 and UKB^12^. PWAS-dup was excluded because of outlying locus size and imprinting mechanism, and 13q12.12-dup was excluded from **d)** owing to the outlying low iPSYCH2015/UKB prevalence ratio. The plots include fitted trend lines (created with geom_smooth). The overall trend across all three psychiatric outcomes for the two locus features and the two CNV prevalence-related features was assessed with generalised estimating equation (GEE) models including all rCNV carriers (Methods), and significant positive associations were found for locus size (P=0.0028) and sum LOEUF score (P=0.0030) as well as the prevalence ratio between iPSYCH2015 and UKB (P=0.0086), but not CNV prevalence in iPSYCH2015 (P=0.31).

To formally test these trends, we used GEE to model the association of these four features with case status across all three diagnosis groups simultaneously among rCNV carriers, while taking full account of the correlation structure among carriers (Methods). In this analysis, both locus size and LOEUF score were significantly associated with overall case prediction (*β*_size_=0.09, P=0.01; *β*_LOEUF_=0.10, P=0.0015) as was the iPSYCH/UKB rCNV prevalence ratio (*β*=0.12, P=0.0086), whereas iPSYCH2015 prevalence was not (*β*=-0.02, P=0.31).

### ID and epilepsy

We applied weighted CPH models to estimate the risk of epilepsy and ID associated with rCNVs in our dataset (Methods). These disorders were not targeted specifically in the iPSYCH2015 case ascertainment and the risk estimates therefore are not population-representative in the same way as for the psychiatric disorders targeted in the study design.

Nine rCNVs were significantly associated with increased risk of both epilepsy and ID (out of 28 and 33 tested, respectively), and further 10 rCNV were associated with increased risk of one or the other disorder. Both deletions and duplications were significantly associated with increased risk of ID at the following loci: 1q21.1, 15q13.3, 16p13.3, 16p11.2, 17q12 and 22q11.2 (Extended Data Figure 2 and Supplementary Table 7).

Three of the nine rCNVs conferring increased risk of both ID and epilepsy (PWAS-dup, 15q13.3-del and 16p13.11-del) were also associated with significantly increased risk for ADHD, ASD and SSD (Extended Data Figure 2, Supplementary Table 3). The greatest increases in risk of both ID (HR=46.45, CI95%: 20.04-107.63) and epilepsy (HR=15.10, CI95%: 7.38-30.90) were associated with PWAS-dup.

## Discussion

The comprehensive assessment of rCNV-associated risk across common psychiatric disorders in an entire population presented in this study, extends beyond current knowledge due to two features of the study design. First, the systematic assessment of deletions and duplications across most known rCNV loci, enabled us to fill a critical gap in the existing literature, especially for ADHD and ASD where our samples are considerably larger and involve more rCNVs than in earlier studies^1,5^. Second, the unbiased population-based study design^23^ allowed accurate revisement of estimates of rCNV prevalence and associated risk of psychiatric disorders.

Our study includes approximately five times as many ASD cases (22,167) and three times as many ADHD cases (26,186) as the largest published rCNV studies^1,5^, and risk estimates for a majority of the examined rCNV are novel for both disorders. We observed six and seven rCNV associated with increased risk of ASD and ADHD, respectively, that had either not been included or not found significant in the previous studies. We confirm high risk of ASD associated with PWAS-dup and 17q12-del, while rCNVs at 16p11.2 were associated with lower increase in ASD risk than previously reported^1^. In general, rCNV-associated increases in risk of ADHD were modest, with HR ranging from 1.4 at 15q11.2 to 4.2 at 17q12. Further, we did not find evidence to support previous reports of high risk of ADHD associated with 22q11.2-del^5^.

Our SSD sample includes 13,126 cases and while some published case-control meta-analyses include larger case samples^6-8^, we provide risk estimates for nine rCNVs not included in those studies^6-8^. We confirm high risk of SSD associated with PWAS-dup (HR=9.2) and moderate SSD-associated risk (HR: 2.0-4.8) with seven other rCNVs. Notably, for six rCNVs our results differ significantly from those of previous studies^6-8^. These include 1q21.1-del, 15q11.2-del, 16p11.2-dup and 17p12-del, for which we find no significant evidence of association with SSD; 22q11.2-del, previously reported with very high risk (as high as OR=68)^6^ but only moderately associated risk (HR=3.3) in our study; and 22q11.2-dup, reported to protect against SSD (OR=0.15)^6^ but here found not to significantly affect SSD risk (HR=1.3, CI95%: 0.70-2.5).

The results for 22q11.2_del are consistent with earlier studies involving Danish registers and biobanks^20-22^, and contrast with several studies that consistently found high risk of SSD associated with 22q11.2-del,^6,17-19^. One explanation that could reconcile this difference is if 22q11.2-del were specifically associated with a late-onset form of SSD, although this hypothesis has not been suggested in past studies of clinically ascertained SSD patients. Also, such a scenario would not explain the risk disparity for 22q11.2-dup. Importantly, we confirm high risk conferred by PWAS-dup of both ASD (HR=21) and SSD (HR=9.2) strongly supporting the validity of the SSD analyses in our study. Earlier studies found risk of ASD^29^ and SSD^30,31^ to be confined to maternally inherited PWAS-dup, but information on parent-of-origin is unfortunately unavailable for rCNV carriers in iPSYCH2015.

Our MDD sample is similar in effective sample size to that of the UKB^11^, which also included many rCNVs assessed in our study. Nonetheless, the UKB study reports high risk for PWAS-dup (OR=8.1) and modest risk increases for four other rCNVs (OR: 1.7-2.7)^11^, while we find no evidence of significantly increased risk of MDD associated with any rCNV. The BPD case sample in our study is small and doesn’t add meaningfully to the limited evidence for rCNV involvement in BPD^10^, while for the broader diagnosis group of “any affective disorder” we find no significant evidence of associated risk among the tested rCNV.

Consistent with previous reports highlighting the pleiotropic effect of rCNVs^1,2^, we observed high pairwise correlation of risk estimates for rCNVs across ASD, ADHD and SSD. Also, our indicative associations for ID and epilepsy mostly overlapped rCNVs conferring increased risk also of ASD, ADHD, and/or SSD. The main exception from this pleiotropic trend was MDD, for which we found no evidence of rCNV-associated risk increase. Also, despite clearly having a pleiotropic effect, the PWAS-dup is associated with substantially higher risk of ASD than of ADHD and SSD. The PWAS locus stands out in several aspects; it is by far the largest of the 18 loci that we could study, and, as mentioned above, has a well-documented parent-of-origin dependent mode of inheritance^29-31^.

While meiotic non-allelic homologous recombination produces *de novo* rCNV deletions more often than duplications^32^, rCNV studies (including this study) consistently report higher population prevalence of duplications^12,26,27^. The general view has been that this prevalence disparity reflects a greater effect on viability and reproductive fitness associated with deletions than duplications, a part of which has been attributed to risk of psychiatric disorders. Our results, however, suggest a more complicated picture.

Firstly, we demonstrate that the deletion-duplication disparity in population prevalence differs widely among rCNV loci, with at least two loci (2q13 and 16p12.1) having a higher prevalence of deletions than duplications, whereas the loci with significantly higher prevalence of duplications include many of the rCNVs most consistently reported to be associated with psychiatric disorders (1q21.1, 15q13.3, 16p11.2 and 22q11.2).

Secondly, we do not observe greater risk associated with deletions than with duplications for the three most relevant psychiatric disorders; ASD, ADHD and SSD. Across the 17 mostly small or medium sized (0.2-1.7 Mb) rCNV loci included in our analysis, both locus size and locus-wide LOEUF scores were independently associated with increased disease risk while rCNV dosage type was not. This is consistent with a recent study that found the summed constraint of genes affected by CNVs to best explain their associated risk with ASD^33^. The same study also reported that risk associated with deletions was attenuated when accounting for IQ, whereas duplication-associated risk was not^33^.

Thirdly, comparison of the population-based rCNV prevalence in our study with that reported for the UKB^12^ found that deletion carriers are specifically underrepresented in the UKB. This suggests that the “healthy volunteer” bias reported in the UKB^13-15^, may in large part derive from cognitive/developmental outcomes (such as IQ and ID/DD/CM) that in turn may follow a different pattern of association with rCNVs than do psychiatric disorders. In fact, in the comparison of risk estimates between our study and published case-control studies (where a similar selection bias is most often present) we also observe more diminished risk for deletions than duplications (with 6/27 vs 2/19 comparisons having significantly lower risk estimates in iPSYCH2015).

Thus, our results point to a disparity between rCNV-associated risk of psychiatric disorders and other developmental/cognitive outcomes, with the latter being more strongly affected by deletions and thereby driving the overall deletion-duplication prevalence disparity. Given that both types of outcomes are selected against in volunteer-based studies such as the UKB, or case-control studies using screened controls, the expectation would be consistent with what we observe; that disparity in rCNV prevalence and associated risk of psychiatric disorders between iPSYCH2015 and UKB/case-control studies is more pronounced for deletions than duplications.

To summarise the above discussion, we propose that; (1) overall, rCNVs are associated with increased risk of psychiatric disorders to a degree that varies according to locus size and gene content but not according to gene dosage; (2) this risk is overall similar across ADHD, ASD and SSD but much less (if at all) pronounced for MDD; (3) some rCNV loci, such as the PWAS locus, may not adhere to these general statements; (4) the overall higher population prevalence of duplications compared to deletions may in large part be driven by larger deletion-associated effects on other (i.e. non-psychiatric) outcomes relevant to viability and/or reproductive fitness (e.g., ID/DD/CM); (5) healthy volunteer bias and use of screened controls has led to an overestimation of rCNV-associated risk of psychiatric disorders and an underestimation of rCNV prevalence in the general population (especially deletions).

The iPSYCH2015 case-cohort, while optimal to accurately assess rCNV-associated risk of its targeted psychiatric disorders, is less well suited to assess risk of other disorders. Also, the relatively young age of participants limits study power for late-onset psychiatric illness, such as BPD. Therefore, we have focussed on the four disorders with the largest case samples (ASD, ADHD, MDD and SSD) and only briefly addressed other disorders (BPD, ID and epilepsy). Also, as several rCNVs are extremely rare, estimates could not be derived for 12 out of 30 rCNVs considered in the study. Finally, while having nation-wide coverage of hospital-based in- and outpatient diagnoses, iPSYCH2015 does not include affected individuals who have been diagnosed and treated solely outside the hospital regimen.

In conclusion, our study provides important insights into the impact of rCNVs on risk of common psychiatric disorders. Population prevalence of rCNVs, especially deletions, is generally higher, and associated risk of psychiatric disorders lower, than previously reported, most drastically so for 22q11.2-del risk of SSD. Notably, we find no evidence of rCNV-associated risk of MDD. In an era where genetics is increasingly being put to use in clinical practice, our results highlight the need for accurate population-based risk estimates for rare genetic exposures such as rCNVs.

## Methods

### Study design

The current study is based on the iPSYCH2015^23^ case-cohort of 140,116 individuals from the 1,657,449 singletons born between May 1, 1981 and December 31, 2008, in Denmark, who were residents in Denmark at 1 year of age and have a mother registered in the Danish Civil Registration System^34^. The case-cohort is made up of two components: (1) Cases: All individuals (n=92,531) who have been clinically diagnosed with major depressive disorder (MDD; ICD10: F32-F33, and ICD-8 296.09, 296.29, 298.09, 300.49, n=37,555), affective disorder (AFF; ICD10: F30-F39, n=40,482), autism spectrum disorder (ASD; ICD10: F84, n=24,975), bipolar disorder (BPD; ICD10: F30-F31, n=3,819), schizophrenia spectrum disorder (SSD; ICD10: F20-F29, n=16,008), schizophrenia (SCZ; ICD10: F20, n=8,113) or attention-deficit/hyperactivity disorder (ADHD; ICD10: F90, n=29,668); according to inpatient and outpatient discharge diagnoses from all Danish hospitals until December 31, 2015, obtained from the Psychiatric Central Research Register (PCRR).^35^ (2) Population comparison Cohort: 50,615 individuals randomly drawn from the same birth cohort as the cases, corresponding to roughly 3% of the entire population in Denmark born in 1981-2008. The total number of cases is less than the sum of the subtotal for each diagnosis, as some individuals have more than one diagnosis; and the total number of samples in the study is smaller than the sum of cases and cohort, as the cohort is a random sample of the population and therefore includes a small number of cases (Extended Data Figure 1).

In addition to the psychiatric diagnosis groups specifically targeted in the iPSYCH2015 case-cohort design, the same information (date of diagnosis) was obtained through PCRR^35^ and the Danish National Patient Registry (DNPR)^36^ for intellectual disability (ID, ICD10: F70-F79; ICD8: 311-315, n=6,969) and epilepsy (ICD10: G40; ICD8: 345 (excl. 345.29), n=4,796).

### Samples and genotyping

For all individuals within the iPSYCH case-cohort, DNA was extracted and whole-genome amplified from available neonatal blood spots retrieved from the Danish Neonatal Screening Biobank (DNSB)^37^, and genotyped with Illumina genotyping arrays (Illumina, San Diego, CA, USA). The iPSYCH2015 case-cohort is an update and expansion of the study base of the iPSYCH2012 case-cohort, described in detail elsewhere^16^. The sampling and genotyping of additional samples (iPSYCH2015i, which when combined with iPSYCH2012 constitute the complete iPSYCH2015 case-cohort) differed in several ways as detailed elsewhere;^23^ most importantly, iPSYCH2015i samples were genotyped using the Global Screening Array v2, whereas iPSYCH2012 samples had been genotyped using the PsychArray V1.0^16^. Single nucleotide polymorphism (SNP) genotype calling and quality control were performed using Illumina’s GenTrain software tool for all samples that could be successfully identified and extracted from DNSB (95.5%). The extraction of B-allele frequency as well as intensity for each probe was performed using Illumina GenomeStudio. Samples with a genotyping call rate lower than 95% or unexplained genotype-estimated sex discordance with the Danish Civil Registration System^35^ were excluded from further analysis.^23^

### CNV calling and quality control analysis

The selection of rCNV loci into this study was largely based on the 54 CNVs studied in the UKB by Crawford *et al*.^12^ with the following exceptions: NRXN1, NPHP1, CRYL1 and CHRNA7, as we considered these either too small to reliably detect in our dataset or to be CNVs of diverse/non-recurrent nature. Also, we did not test duplications spanning BP3-BP5 on 15q11-q13 specifically, but instead applied a set of hierarchical rules to deal with rCNVs overlapping more than one adjacent recurrent CNV loci (Supplementary Note 2). Thus, we defined 30 distinct recurrent CNV loci, in which we searched for both deletions and duplications spanning at least ⅔ of the defined boundaries, as further outlined in Supplementary Table 1 and in Supplementary Note 2.

Processing and filtering of PennCNV^24^ calls was done using in-house developed R-package QCtreeCNV,^25^ and the resulting putative calls visually inspected using in-house developed graphical interface DeepEye.^25^ To filter samples of poor quality we applied similar threshold values for the main sample-wide quality measures as applied in other large studies, e.g. of the UKB^12^; LRR-SD ≥ 0.35, BAF-drift ≥ 0.005, and |GCWF| ≥ 0.02 (Extended Data Figure 4), which removed 3,130 samples. We assessed the quality of the visual inspection of the 11,862 inspected calls through intra- and inter-rater reliability (IRR, Supplementary Table 1) and tested whether the latter, as well as the fraction of calls where carrier status could not be determined (“unknown”), correlated with LRR-SD and/or with locus, array type or CNV type (Supplementary Table 8). We then performed a series of analyses to determine whether sensitivity in CNV detection and/or specificity of visual validation of putative CNV calls was affected by measurement noise (Supplementary Table 9) or genotyping array (Supplementary Table 10). The details of these analyses and their results are provided in Supplementary Note 2.

### Statistical analysis

#### Estimating and comparing rCNV prevalence

We calculated population-based rCNV prevalence (with CI95%) from the full iPSYCH2015 case-cohort using the *svydesign()* and *svyciprop()* functions from the *survey*^38^ package in R, with finite population correction (fpc) to account for oversampling of cases. Briefly, we divided the post-QC number of (a) case individuals (79,535) and (b) individuals from the randomly drawn population subcohort (43,311) with the total number of corresponding individuals in the source population (92,531 and 1,657,449) to derive the sampled population fractions; 0.85955 and 0.02613, respectively. Samples from overlapping individuals (cases-in-subcohort) were assigned the case population fraction (0.85955). This resulted in an effective population-based sample size corresponding to 45,609 individuals. We compared the overall and per-locus prevalence of deletions and duplications with a Welch’s test of difference between two measures assuming unequal variance. Briefly, we defined the difference; *d = abs(log(p_DEL_/p_DUP_))*, the standard error of the difference; *SEd = √(SE ^2^+SE_DUP_ ^2^)*, and the p-value; *P = 2*(1-pnorm(d/SEd))*, where *p_DEL_* and *SE_DEL_*, and *p_DUP_*and *SE_DUP_*, indicate the prevalence and standard error of prevalence retrieved with *svyciprop()* for deletions and duplications, respectively. We calculated rCNV prevalence in the UKB directly from carrier counts provided in Crawford *et al*.^12^ and CI95% calculated as follows: *CI95% = qbeta(c(0.05/2,1-0.05/2), nCarrier+0.5, nTotal-nCarrier+0.5)*, where *nCarrier* and *nTotal* indicate the number of carriers of the rCNV and the total number of assessed samples (421,268), respectively. We then compared the rCNV prevalence in iPSYCH2015 and UKB with a Welch’s test, as described above. P-values were in both instances adjusted for multiple comparisons with a false discovery rate (FDR) method, using the *p.adjust* function of the (default) *stats* package in R.

#### Estimating rCNV-associated risk of iPSYCH disorders with survival analysis

We used weighted Cox proportional hazard (CPH) models from the *survival*^39^ package in R to estimate the rCNV-associated risk of the five psychiatric disorders targeted by the iPSYCH2015 case-cohort design (ADHD, ASD, BPD, MDD and SSD), as well as of the broader diagnosis groups; any affective disorder and any iPSYCH disorder, and the narrower (compared to SSD) diagnosis of schizophrenia (ICD10; F20). The outcome in the CPH models was the age at first hospital diagnosis with the index disorder, age at censoring or age at death - whichever came first - and the exposure was carrier status (i.e. having deletion or duplication, respectively, versus normal copy number) for the rCNV. All models were sex-stratified. Subjects were censored if they had not received an index diagnosis by the end of the follow-up period (December 31st 2015), emigrated or otherwise had been lost to follow-up. To obtain unbiased population-based estimates, the inverse probability of sampling (IPS) weights were used as introduced by Barlow *et al*.^40^ The SEs of regression coefficients were computed by a robust estimator to derive CI95% and test for the significance of rCNV-associated HRs.

#### Estimating omnibus rCNV-associated risk of the four main iPSYCH2015 disorders

We used generalised linear models (GLMs) to assess the overall effect of rCNVs on the prediction of each of the four main iPSYCH2015 disorders (ADHD, ASD, MDD and SSD) separately. For this analysis we defined a categorical variable with separate levels for each rCNV and “no rCNV” as reference (individuals carrying more than one rCNV were assigned the level corresponding to the rCNV with a higher risk estimate for “any iPSYCH2015 disorder”). Then for each disorder, a full model including this categorical rCNV carrier status as an independent variable in addition to AEF and biological sex (as determined at birth), was compared with a nested model including only AEF and biological sex. Both models included only individuals either diagnosed with the respective diagnosis or belonging to the random population subcohort. A likelihood ratio test (LRT) was used to test the omnibus rCNV effect by comparing the full model with the nested one.

#### Comparison of rCNV-associated risk across diagnoses

We implemented generalised estimating equation (GEE) models using *glmgee()* function from *glmtoolbox*^41^ in R to test for overall and/or rCNV-specific differences in associated risk between pairs of diagnoses (ASD, ADHD and SSD). We limited each analysis to those individuals who were part of the random population subcohort or diagnosed with either of the two compared diagnoses, and allocated two lines to each sample ID in the data structure to account for the possibility of having been diagnosed with both disorders. Thus, the binary case outcome in the model was independent between diagnoses for each included study subject. For each pairwise comparison, two GEE models (full and nested) were constructed, with case status as outcome and clustering on sample ID. Independent categorical variables were rCNV and diagnosis (i.e. indicating which of the two compared disorders each line’s outcome value belonged to for each study subject, in each comparison the diagnosis with a larger case sample size was assigned as reference level), along with sex and AEF. In the analysis of overall rCNV-associated differences between diagnoses, rCNV status was defined as binary carrier status for “any rCNV”, while in the analysis of rCNV-specific cross-diagnosis differences, a categorical variable with separate levels for each rCNV was specified for rCNV status with “no rCNV” as the reference (if individuals carried more than one rCNV, we assigned them the level of the rCNV with higher associated estimate for “any iPSYCH disorder”). In addition to the variables specified above, the full models included an interaction term between rCNV status (binary or categorical) and diagnosis, whereas the nested models did not. We then used anova test from the *glmtoolbox* package (with *test=”score”* and *varest=”model”*) to compare each full model with its respective nested model. Furthermore, to extract significant coefficient of interaction between rCNVs and the diagnosis, we performed *post hoc* analysis on summary results of the GEE models (i.e the model that included the interaction term) by applying FDR correction on the p-values corresponding to all coefficients in the model outputs.

#### Analysis of locus type effect on rCNV-associated risk across the diagnoses

To investigate the rCNV dosage type effect (i.e., deletion v. duplication) on rCNV pathogenicity across ASD, ADHD, and SSD, we again applied GEE models similarly as described above, although only among rCNV carriers. To account for multi-diagnoses individuals, three rows were assigned to each individual id in the data structure corresponding to the three included diagnoses, and the binary case status was defined in each row independently based on whether the rCNV carrier had any of the diagnoses. The GEE models included diagnosis, rCNV type (del v. dup), and locus of the corresponding rCNV carrier as independent categorical variables, to predict the case status as the model outcome (binary variable), while adjusting for AEF and sex and clustering on ids. The reference categories for rCNV type, locus, and diagnosis as categorical variables were set as deletion, 15q11.2, and ADHD respectively in each model. To evaluate overall locus type effect across three diagnoses, we compared the full model with a nested model excluding rCNV type from the covariates by performing ANOVA test from the same package and default mentioned above. Additionally, to test locus-specific rCNV type effects, we built another model containing an interaction between rCNV type and locus and compared it with the base model using ANOVA test.

#### Analysis of locus features and rCNV prevalence as predictors of penetrance

For analysis of locus features’ effect on rCNV pathogenicity, we constructed two GEE models among all rCNV carriers. The first model contained LOEUF score and locus size, while the second model contained both rCNV iPSYCH prevalence and iPSYCH/UKB prevalence ratio as independent variables to predict the case status (binary outcome) separately. Both models included AEF and sex as covariates and sample ID as clustering variable. In both models the test variables (LOEUF score and size, and iPSYCH prevalence and iPSYCH/UKB prevalence ratio, respectively) were log-transformed.

#### Estimating rCNV-associated risk of ID and epilepsy with survival analysis

To study ID and epilepsy, we fitted weighted CPH models with age at the first hospital diagnosis (otherwise censoring date) as the primary outcome on the entire case-cohort sample. Considering the frequent comorbidity between ID and epilepsy with psychiatric outcomes, we assigned IPS weights to the individuals depending on whether they belonged to the case or cohort component of iPSYCH2015 in a similar way as for the primary outcomes^40^. All the analyses included sex as the stratification variable and rCNV status as the independent variable, which was coded as described in primary analyses of iPSYCH outcomes.

#### Sensitivity analyses using generalised linear models for all the studied outcomes

Sensitivity analysis was performed utilising generalised linear models (GLMs) in parallel to CPH models to compute rCNV-associated ORs for all the studied outcomes. When studying psychiatric diagnoses, only the individuals who had the corresponding outcome or were in the random cohort were included in the analyses, with the diagnosis as the binary outcome and rCNV as independent variable in each model accounting for AEF and sex of the individuals and genotyping array (i.e whether individual was genotyped on PsychArray or GSA). However, for ID and epilepsy, GLMs were fitted on the entire case cohort, thus accounting for the 5 main iPSYCH outcomes, AEF and sex of participants and genotypic array.

All statistical analyses were conducted in R^42^, version 4.2.3.

## Data Availability

"All data produced in the present study (other than sensitive person-level data, which by requirement of the data custodian and Danish legislation
cannot be shared; i.e data that corresponds to fewer than 5 individuals) is available upon reasonable request to the corresponding author(s)."

