## Supplementary text for "Population-based Risk of Psychiatric Disorders Associated with Recurrent CNVs"

**Population-based Risk of Psychiatric Disorders Associated with Recurrent CNVs: Supplementary Notes, Extended Data Figures and Supplementary Tables**

Supplementary Note 1	page 1

Supplementary Note 2	page 2

Extended Data Figure 1	page 5

Extended Data Figure 2	page 6

Extended Data Figure 3	page 7

Extended Data Figure 4	page 8

Supplementary Table 1	page 9

Supplementary Table 2	page 10

Supplementary Table 3	page 11

Supplementary Table 4	page 13

Supplementary Table 5	page 14

Supplementary Table 6	page 14

Supplementary Table 7	page 15

Supplementary Table 8	page 16

Supplementary Table 9	page 17

Supplementary Table 10	page 18

**Supplementary Note 1**

**iPSYCH investigators**

The building of the vast research data source underlying the iPSYCH2015 case-cohort involved many more researchers than those who were directly involved with the study of rCNVs described in this manuscript. We have therefore included those researchers who played a crucial role in building and maintaining the iPSYCH2015 dataset under the banner of the "iPSYCH Investigators" group authorship. Their names and affiliations are as follows:

Anders D. Børglum^1-3^, David M. Hougaard^4^, Merete Nordentoft^5^, Ole Mors^6^, Preben B. Mortensen^2,7-9^, Thomas Werge^2,10-12^, Jakob Grove^1-3,13^, Thomas D. Als^1-3^, Alfonso Buil^10,11^, Anders Rosengren^10^, Andrés Ingason^10,11^, Andrew J. Schork^10,11^, Dorte Helenius^10^, Jesper Gådin^10^, Richard Zetterberg^10^, Vivek Appadurai^10^, Joeri Meijsen^10^, Kajsa-Lotta Georgii Hellberg^10^, Bjarni J. Vilhjálmsson^7,13^, Carsten B. Pedersen^7^, Esben Agerbo^7^, Jakob Christensen^7^, Liselotte V. Petersen^7^, Marianne Giørtz Pedersen^7^, Jonas Bybjerg-Grauholm^4^, Marie Bækvad-Hansen^4^

1. ^Department of Biomedicine, Aarhus University, Aarhus, Denmark.^
2. ^The Lundbeck Foundation Initiative for Integrative Psychiatric Research, iPSYCH, Denmark.^
3. ^Center for Genomics and Personalized Medicine, Aarhus, Denmark.^
4. ^Department for Congenital Disorders, Statens Serum Institute , Copenhagen, Denmark.^
5. ^Mental Health Centre Copenhagen, Capital Region of Denmark, Copenhagen University Hospital, Copenhagen, Denmark.^
6. ^Psychosis Research Unit, Aarhus University Hospital-Psychiatry, Denmark.^
7. ^NCRR - National Centre for Register-Based Research, Business and Social Sciences, Aarhus University, Aarhus V, Denmark.^
8. ^Centre for Integrated Register-based Research, CIRRAU, Aarhus University, Aarhus, Denmark.^
9. ^Centre for Integrative Sequencing, Department of Biomedicine and iSEQ, Aarhus University, Aarhus, Denmark.^
10. ^Institute of Biological Psychiatry, Mental Health Services, Copenhagen University Hospital, Copenhagen, Denmark.^
11. ^Lundbeck Foundation Center for GeoGenetics, GLOBE Institute, University of Copenhagen, Copenhagen, Denmark.^
12. ^Department of Clinical Medicine, University of Copenhagen, Copenhagen, Denmark.^
13. ^BiRC, Bioinformatics Research Centre, Aarhus University, Aarhus, Denmark.^

**Supplementary Note 2**

**CNV calling, filtering, visual inspection, and quality control analysis**

Intensity file preprocessing and CNV calling

Raw genotype data in the form of Illumina intensity files were stored on a secure partition on the HPC cluster GenomeDK (<https://genome.au.dk/>). We retrieved a total of 128,225 intensity files that could be used in the CNV calling pipeline. A small subset of samples had been genotyped more than once, and for these samples we only kept PennCNV^1^ calls from the intensity file with lower log-R-ratio standard deviation (LRR-SD; see below). In total, 123,377 unique iPSYCH2015 samples from the case-cohort design (Methods) were available for CNV analysis. To reduce probe intensity noise and erroneous CNV calling, we filtered the intensity files from each genotyping array type separately to include only biallelic autosomal SNPs mapping uniquely to the Haplotype Reference Consortium (HRC)^2^ hg19 reference map, with a minor allele frequency of at least 0.1%, which yielded 280,700 and 509,754 probes for the PsychArray and GSA, respectively. Next, CNV calling was performed using PennCNV^1^ in batches of 1,200 samples, using Population Frequency of B allele (PFB) files based on the actual B allele frequencies (BAF) in each batch. We used the default hidden Markov model (HMM) file provided by PennCNV^1^ and produced array-specific GC-content (GCC) files based on the file “hg19.gc5Base.txt.gz” downloaded from the Genome browser of the University of California in Santa Cruz (<https://genome.ucsc.edu/>). Initial PennCNV calls were obtained with the script “*detect_cnv.pl*” setting minimum number of probes (--minsnp) at 5, and minimum length (--minlength) at 1000 bp. We then merged adjacent calls, with the PennCNV script “*clean_cnv.pl*” using the settings “*--fraction 0.2 --bp*” whereby two calls are merged if the gap between them corresponds to less than 20% of the combined length of the calls in terms of base pairs.

Subsetting and filtering CNVs at target recurrent CNV (rCNV) loci

PennCNV calls and sample QC files (containing LRR- and BAF-derived measures for each sample informing about overall sample quality) were imported into *R*, version 4.0.5. Subsetting and filtering of calls for the current study was then performed using the in-house developed *R* package *QCtreeCNV* (<https://github.com/SinomeM/QctreeCNV>). A general overview of the filtering steps and the *QCtreeCNV* pipeline is provided elsewhere^3^. First, we selected the calls in the 30 target autosomal loci (Supplementary Table 1) and merged remaining adjacent calls in each sample using the function “*QCtreeCNV::select_stitch_calls()*” with default values (minsnp = 20, maxgap = 0.5, minoverlap = 0.2) for all loci except WBS and 22q11.2 where required minimum overlap was increased to 0.35. Also, the required minimum number of probes was reduced from 20 to 15 at four loci (TAR, 2q21.1, 16p12.1 and 16p11.2d) after we found upon initial visual inspection that a substantial fraction of true calls at these loci (ranging from 5% for 16p12.1 to nearly half of true 16p11.2d calls) involved 20-24 probes. We then used the function “*QCtreeCNV::qctree()*” to select putative carriers to visually inspect, using default values for all parameters except “*st5maxlogr1=0.55*”.

Sample filtering and visual inspection

The distribution of per-sample quality measures; standard deviation of LRR (LRR-SD), BAF-drift, and GC-wave factor (GCWF), is shown in Extended Data Figure 4. We discarded samples exceeding any of the three following threshold values; LRR-SD ≥ 0.35, BAF-drift ≥ 0.005 and/or |GCWF| ≥ 0.02. This resulted in the removal of 3,130 samples, corresponding to 2.5%. After removal of these samples and the filtering of rCNV calls as described above, we plotted LRR and BAF values for each of the 12,074 remaining rCNV calls. Each plot was then inspected by the senior author twice (to derive intra-rater reliability estimates) and by at least one of the two primary analysts, using *DeepEye*, an in-house developed graphical interface, designed to facilitate fast rating of CNV calls and accurate storage thereof, described in detail elsewhere^3^. Each call was rated as either true (T), false (F) or unknown (U). All calls with disagreement across the 3-4 ratings of the 2-3 raters were rated again jointly by all three raters to reach a consensus. We considered as true, those calls that had a LRR and BAF pattern consistent with a deletion or duplication (or, in some instances, triplication) overlapping at least 2/3 of the locus in question, irrespective of the actual boundaries of the putative PennCNV calls.

CNV grouping and initial quality control

We split the 30 loci into four groups depending on locus size, while keeping the 15q11.2 locus as a separate fifth group, as rCNVs at this locus are by far the most prevalent, both among deletions and duplications (for the locus size ranges see Supplementary Table 1). Some of the loci are in close proximity to another locus or loci (Supplementary Table 1) and many true rCNV calls were found to span more than one locus. To account for this, we applied hierarchical filtering of true calls by removing (only in instances involving same rCNV type): (a) true 15q11.2, 15q11.2d, 15q13.1 and 15q13.3 calls in samples with a true Prader-Willi/Angelman Syndrome (PWAS) call; (b) true 22q11.2b and 22q11.2d calls in samples with a true 22q11.2 call; (c) true 16p12.1 and 16p11.2d calls in samples with a true 16p11.2 call; (d) true TAR calls in samples with a true 1q21.1 call; (e) true 7q11.23d calls in samples with a true WBS call; (f) true 15q11.2 calls with a true 15q11.2d call; (g) true 15q13.1 calls with a true 15q13.3 call; and (h) true 16p12.3 calls with a true 16p13.1 call (in total 371 redundant calls were removed).

We then applied rigorous quality control to verify the accuracy and validity of the rCNV calling and subsequent visual inspection. First, we derived the fraction of true calls at each locus, and removed three loci, with <5% of calls deemed true; 15q24, 7q11.23d and WBS (with true call fraction ranging from 0% to 1.2%, Supplementary Table 1). Of the remaining 7,373 calls, 3,559 were found to be true, corresponding to 48.3%, a fraction which varied substantially across loci, from 6.2% (29 of 471) at 3q29 to 100% at 16p12.3 and 17p12 (Supplementary Table 1).

Next, we plotted the rates of T, F and U calls as well as the inter-rater reliability (IRR) across increasing LRR-SD intervals by rCNV type, locus group and genotyping array (Supplementary Table 8). Unsurprisingly, the fraction of calls rated as unknown (U) increased with increased LRR-SD, while the IRR decreased with increased LRR-SD, with both trends most noticeably observed for the small (S) and medium-sized (M) loci. This indicates reduced confidence in our rating of calls in (a) samples with higher compared to lower LRR-SD, and (b) smaller compared to larger loci. Also, the fraction of unknown calls and the IRR differed significantly between genotyping arrays (see Supplementary Table 8).

Population-based rCNV prevalence by LRR-SD and genotyping array

To study whether this association between (a) LRR-SD, locus size and genotyping array, and (b) confidence in initial rating of calls had affected the validity of our final consensus ratings, we tested for association between these variables and the population-based prevalence of true calls (i.e. calls that we consider true based on our visual inspection). The analyses were done in *R* using the *survey* package^4^ functions *svydesign()*, *svyciprop()* and *svyglm()*, and applying finite population correction (fpc) to account for oversampling of cases in the case-cohort sample. Essentially, we followed the same approach as outlined in (Methods), while the *svyglm()* was used to assess the potential effect of LRR-SD (ranging from 0.0956-0.3499 across samples) and/or genotyping array (PA or GSA). Neither the overall rCNV prevalence nor that of any subclass based on CNV type, genotyping array or locus size, was associated with LRR-SD (P>0.05 for all tests, Supplementary Table 9, note that while prevalence is shown for bins of increasing LRR-SD, the test for LRR-SD effect was done based on per-sample values for all 120,247 samples). Also, neither overall rCNV prevalence nor that of rCNVs at any of the 18 loci specifically, differed significantly between samples genotyped on the PsychArray and GSA, neither overall (P>0.05 for all tests, Supplementary Table 10).

**Extended Data Figures**

Extended Data Figure 1

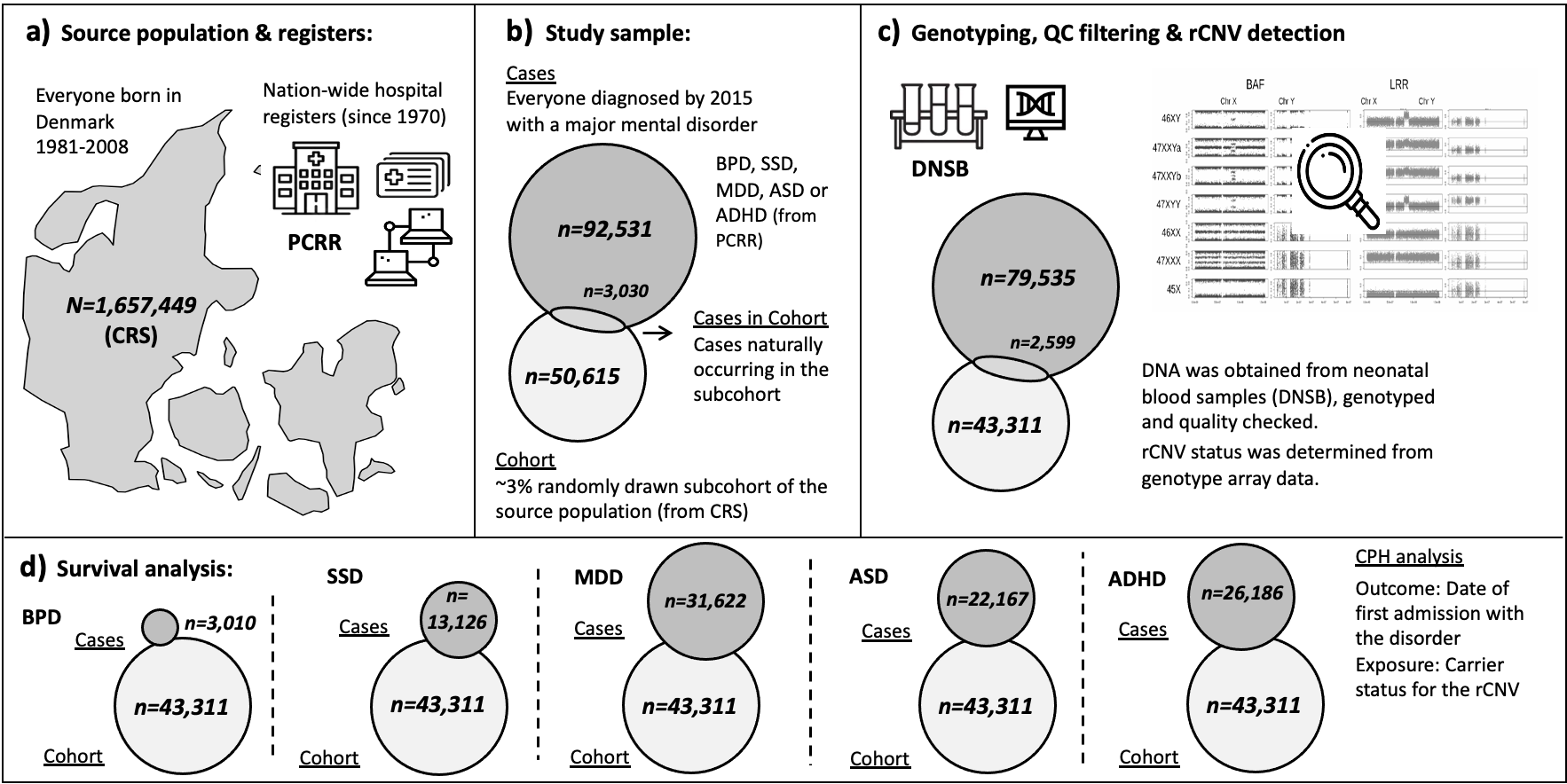

***The iPSYCH2015****^23^* ***Case-Cohort design at a glance.*** ***a)*** *The source population is everyone born in Denmark between May 1st, 1981 and Dec 31st, 2008 to a mother registered in the Civil Registration System (CRS)^34^ and residing in Denmark at their first birthday. The Psychiatric Central Research Register (PCRR)^35^ holds records on diagnoses from all psychiatric departments and mental hospitals in Denmark since 1970.* ***b)*** *The study design included everyone from the source population who had been diagnosed with any of the following index psychiatric disorders; attention-deficit hyperactivity disorder (ADHD), autism spectrum disorder (ASD), major depressive disorder (MDD), schizophrenia spectrum disorder (SSD) or bipolar disorder (BPD) by Dec 31st, 2015 (****Cases****); as well as a subcohort, randomly drawn from the source population (****Cohort****), which by necessity includes some Case individuals (Cases-in-Cohort).* ***c)*** *Blood samples were retrieved from the Danish Neonatal Screening Biobank (DNSB)^37^, and, following DNA extraction, microarray genotyping and initial quality control around 85% of samples from the initial study design remained. Copy number variants (CNVs) were called by PennCNV^24^ and carrier status at 30 recurrent CNV loci (rCNV) verified by our researchers using the in-house developed QC-tree protocol^25^.* ***d)*** *Population-valid risk estimates for the five index psychiatric disorders associated with carriage of each rCNV were assessed by Cox Proportional Hazard (CPH) function with age at first hospitalisation with the respective disorder as outcome, accounting for date of birth and biological sex, and applying inverse probability of sampling (IPS) weights^40^.*

Extended Data Figure 2

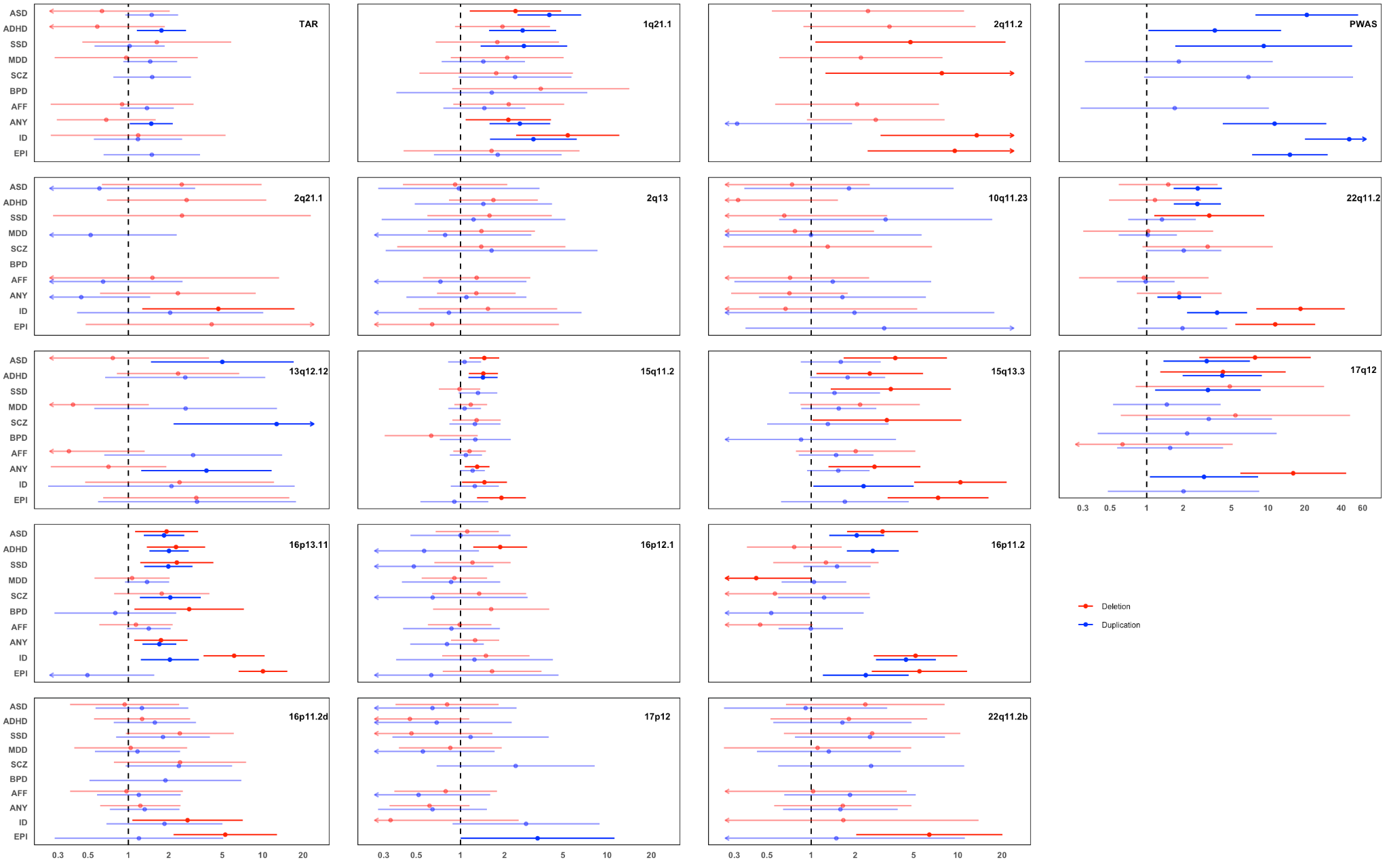

***rCNV-associated risk of psychiatric diagnoses, Any iPSYCH disorder, ID and epilepsy in IPSYCH2015 case-cohort at each studied locus:*** *rCNV-associated Hazard ratios (HR) and 95% confidence intervals (CI95%) for all the included diagnoses are demonstrated per locus separately across 18 loci. HRs and 95% confidence intervals (error bars) for deletions and duplications are indicated by red and blue colour, respectively, and bolded in case of having P<0.05 (i.e. with CI95% not overlapping HR=1). Note that range of values the on x-axes (shown in logarithmic scale) differs between the first three rows and the last row of the plots. Abbreviations for all the diagnoses denoted on the plots are as follows: ADHD; attention-deficit hyperactivity disorder, ASD; autism spectrum disorder, MDD; major depressive disorder, SSD; schizophrenia spectrum disorder, SCZ; schizophrenia, BPD; bipolar disorder, AFF; any affective disorder, ANY; any iPSYCH disorder, ID; intellectual disability, and EPI; epilepsy. Any iPSYCH disorder is defined as being diagnosed with any of index disorders in iPSYCH2015 case-cohort.*

Extended Data Figure 3

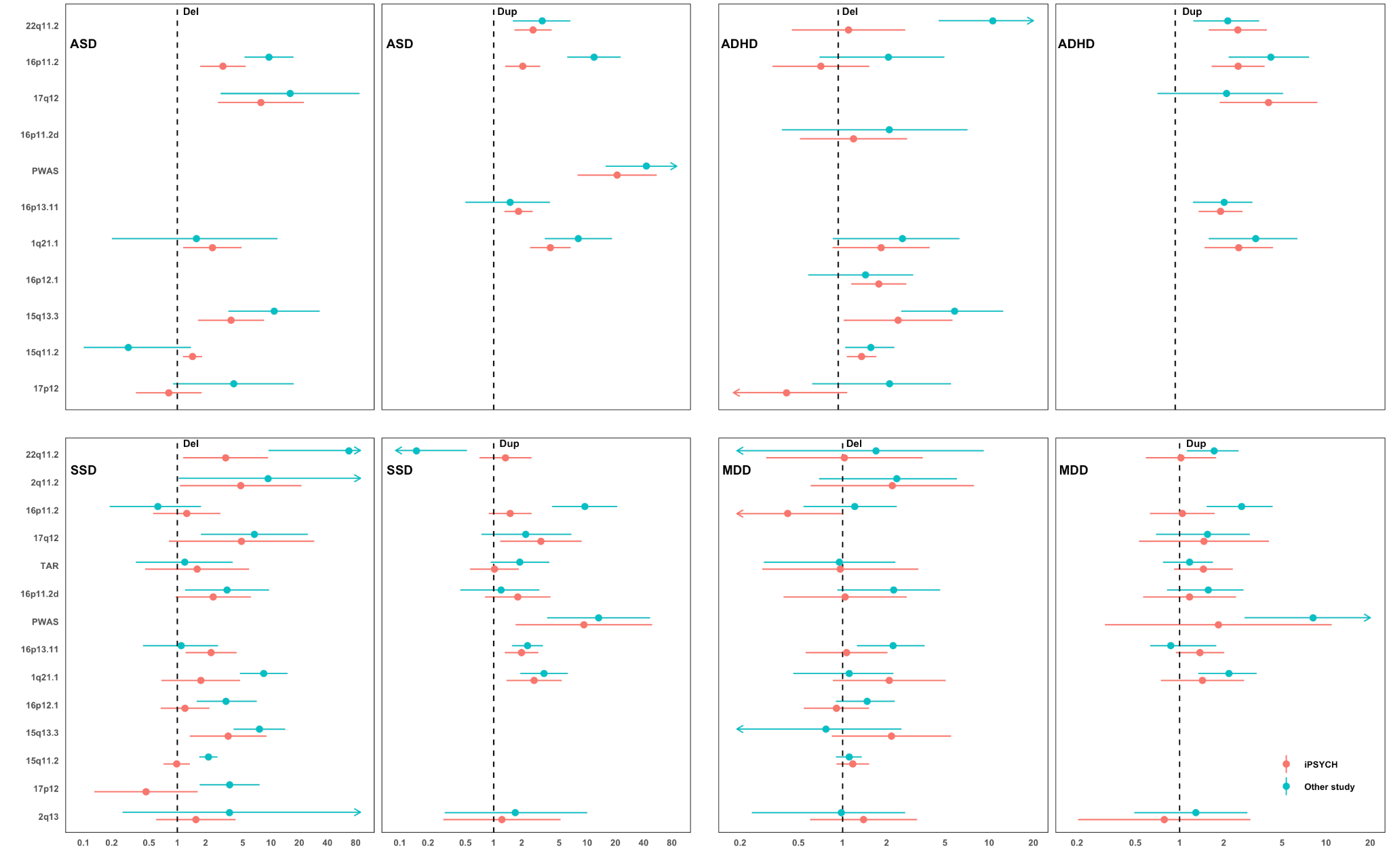

***Comparison of rCNV-associated risk of psychiatric disorders in iPSYCH2015 case-cohort with previously reported risk estimates from case-control studies.*** *We compared the population-valid risk of ASD, ADHD, SSD and MDD estimated for iPSYCH2015 (red) with previously published risk estimates (blue) associated with rCNVs across the 18 loci assessed in this study. The number of rCNVs with available published risk estimates ranged from 11 for ASD1 to 13 for ADHD4, 20 for MDD10 and 22 for SSD5-7 (Supplementary Table 4). Risk estimates for iPSYCH2015 are hazard ratios (HR), while those for the compared case-control studies are odds ratios (OR). Error bars indicate 95% confidence intervals. The x-axis is log-scaled and differs between the two columns. Note the loci shown on y-axis varies between the first row(ASD, ADHD) and the second row (SSD, MDD) and the values on x-axis are log transformed. Only HRs with comparable ORs from other studies are displayed on the plots. ASD: autism spectrum disorder, ADHD: attention-deficit/hyperactivity disorder, SSD: schizophrenia spectrum disorder, MDD: major depressive disorder.*

Extended Data Figure 4

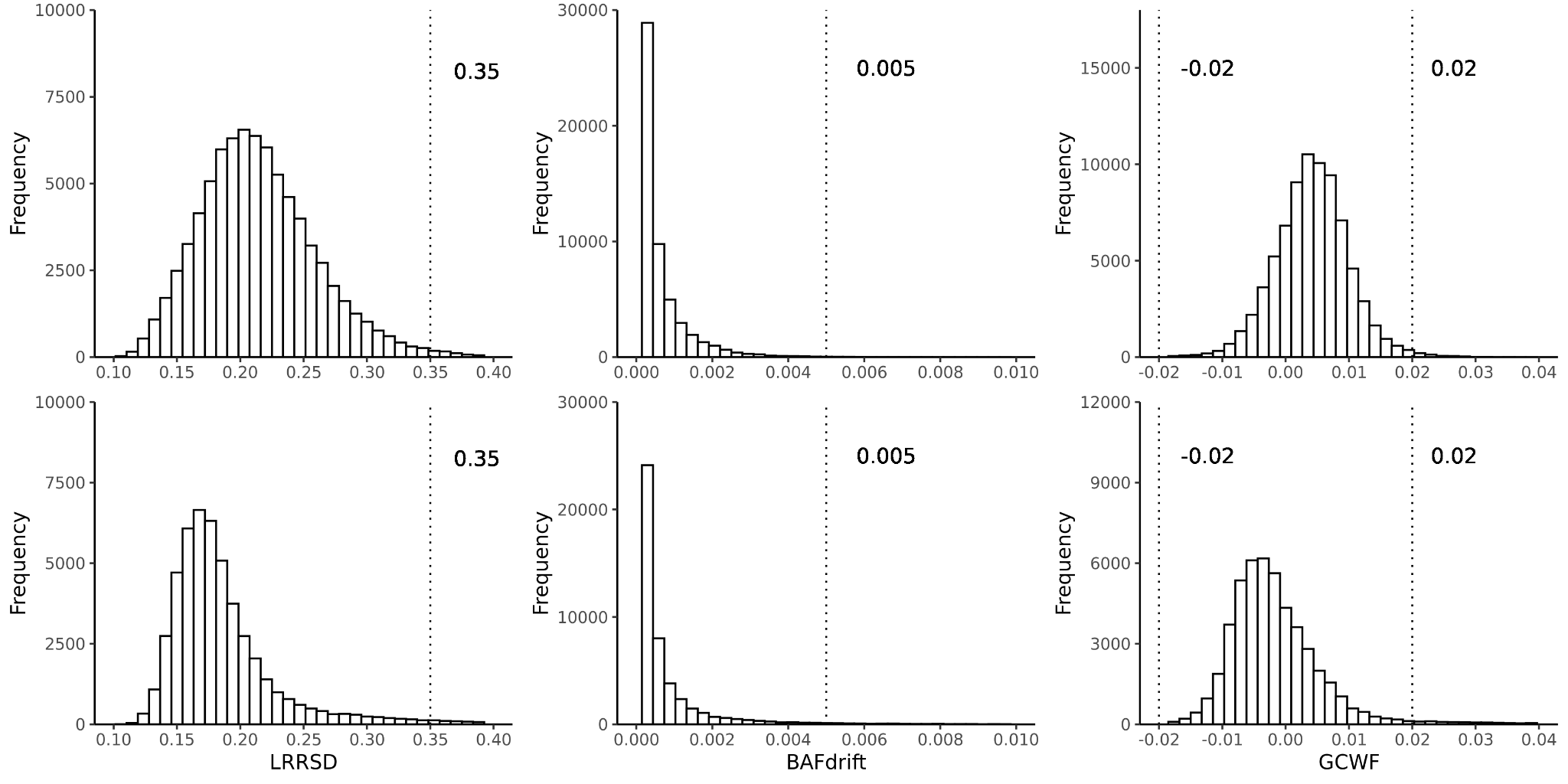

**Per-sample CNV-calling quality measures.** Distribution of per-sample LRR-SD (left), BAF-drift (middle) and GCWF (right) in iPSYCH2015 (Upper row: Samples genotyped with Illumina PsychArray in the initial iPSYCH2012 study^16^. Lower row: Samples genotyped with Illumina GSA in the iPSYCH2015i expansion, which together with iPSYCH2012 samples constitutes iPSYCH2015^23^). Vertical lines and adjacent numeric labels indicate the cutoff values for each measure used to discard outlying samples with intensity data of too low quality to reliably call rCNVs. Samples falling outside of any cutoff value were excluded.

**Supplementary tables**

*Supplementary table 1: rCNV loci selected for study in the iPSYCH2015 case-cohort sample*

| **rCNV locus** | **Hg19 position (Mb)** | **Size (Mb)** | **Locus group^a^** | **n Probes (PA/GSA)^b^** | **Genes (n)^c^** | **LOEUF (sum^-1^)^d^** | **Inspected Calls (n)^e^** | **True Calls (n)^e^** | **True Calls (%)** | **Intra-RR (TvF\|U)^f^** | **Inter-RR (TvF\|U)^f^** | **Filtered in QC^g^** |
| --- | --- | --- | --- | --- | --- | --- | --- | --- | --- | --- | --- | --- |
| TAR | Chr1:145.39-145.81 | 0.42 | S | 55/56 | 16 | 22.2 | 269 | 200 | 74.3% | 97% | 95% |  |
| 1q21.1 | Chr1:146.53-147.39 | 0.86 | M | 135/177 | 7 | 9.1 | 213 | 202 | 94.8% | 100% | 99% |  |
| 2q11.2 | Chr2:96.74-97.68 | 0.94 | M | 82/108 | 18 | 43.8 | 339 | 23 | 6.8% | 99% | 85% |  |
| 2q13 | Chr2:111.39-112.01 | 0.62 | M | 71/131 | 3 | 5.0 | 87 | 82 | 94.3% | 100% | 97% |  |
| 2q21.1 | Chr2:131.48-131.93 | 0.45 | S | 31/68 | 4 | 5.1 | 55 | 32 | 58.2% | 93% | 86% |  |
| 3q29 | Chr3:195.72-197.35 | 1.63 | L | 286/341 | 21 | 41.3 | 471 | 29 | 6.2% | 100% | 100% | excl.^3^ |
| WBS | Chr7:72.74-74.14 | 1.4 | L | 118/189 | 23 | 50.2 | 1,872 | 24 | 1.3% | 100% | 100% | excl.^1^ |
| 7q11.23d | Chr7:75.14-76.06 | 0.92 | M | 93/168 | 12 | 17.0 | 2,441 | 6 | 0.2% | 100% | 99% | excl.^1^ |
| 8p23.1 | Chr8:8.10-11.87 | 3.77 | XL | 673/1,026 | 27 | 27.5 | 11 | 8 | 72.7% | 100% | 100% | excl.^2^ |
| 10q11.23 | Chr10:49.39-51.06 | 1.67 | L | 287/345 | 17 | 23.5 | 47 | 36 | 76.6% | 100% | 98% |  |
| 10q23 | Chr10:82.05-88.93 | 6.88 | XL | 660/1,165 | 24 | 39.9 | 9 | 8 | 88.9% | 100% | 100% | excl.^2^ |
| 13q12.12 | Chr13:23.56-24.88 | 1.32 | L | 273/403 | 6 | 8.0 | 41 | 40 | 97.6% | 100% | 98% |  |
| 15q11.2 | Chr15:22.81-23.09 | 0.28 | 15q11.2 | 49/66 | 4 | 8.4 | 1,253 | 1,188 | 94.8% | 99% | 98% |  |
| 15q11.2d | Chr15:23.68-24.33 | 0.65 | M | 91/143 | 4 | 7.3 | 11 | 10 | 90.9% | 100% | 100% | excl.^2^ |
| PWAS | Chr15:24.82-28.39 | 3.57 | XL | 471/871 | 8 | 19.0 | 57 | 54 | 94.7% | 100% | 98% |  |
| 15q13.1 | Chr15:29.16-30.38 | 1.22 | L | 127/253 | 4 | 19.1 | 25 | 22 | 88.0% | 100% | 100% | excl.^3^ |
| 15q13.3 | Chr15:31.08-32.46 | 1.38 | L | 153/253 | 6 | 8.7 | 307 | 168 | 54.7% | 100% | 99% |  |
| 15q24 | Chr15:72.90-78.15 | 5.25 | XL | 460/701 | 64 | 112.3 | 17 | 0 | 0.0% | 100% | 100% | excl.^1^ |
| 16p13.11 | Chr16:15.51-16.29 | 0.78 | M | 125/213 | 8 | 19.0 | 500 | 423 | 84.6% | 100% | 98% |  |
| 16p12.3 | Chr16:16.86-18.17 | 1.31 | L | 146/316 | 1 | 3.0 | 10 | 10 | 100.0% | 100% | 100% | excl.^2^ |
| 16p12.1 | Chr16:21.95-22.43 | 0.48 | S | 47/50 | 8 | 9.0 | 287 | 210 | 73.2% | 94% | 94% |  |
| 16p11.2d | Chr16:28.82-29.05 | 0.23 | S | 22/35 | 9 | 21.4 | 201 | 116 | 57.7% | 92% | 87% |  |
| 16p11.2 | Chr16:29.65-30.20 | 0.55 | S | 50/53 | 28 | 42.9 | 920 | 266 | 28.9% | 98% | 92% |  |
| 17p12 | Chr17:14.14-15.43 | 1.29 | L | 237/406 | 6 | 6.5 | 69 | 69 | 100.0% | 100% | 99% |  |
| PLS | Chr17:16.81-20.21 | 3.4 | XL | 293/444 | 49 | 76.6 | 144 | 10 | 6.9% | 100% | 99% | excl.^2^ |
| 17q11.2 | Chr17:29.12-30.27 | 1.15 | M | 101/154 | 14 | 26.3 | 27 | 8 | 29.6% | 100% | 100% | excl.^2^ |
| 17q12 | Chr17:34.81-36.22 | 1.41 | L | 161/282 | 15 | 37.1 | 94 | 93 | 98.9% | 100% | 98% |  |
| 22q11.2 | Chr22:18.90-20.30 | 1.4 | L | 172/328 | 27 | 50.4 | 1,689 | 192 | 11.4% | 100% | 100% |  |
| 22q11.2b | Chr22:20,71-21,47 | 0.76 | M | 86/141 | 15 | 24.2 | 141 | 52 | 36.9% | 97% | 97% |  |
| 22q11.2d | Chr22:21,92-23,65 | 1.73 | L | 166/287 | 20 | 30.6 | 96 | 8 | 8.3% | 100% | 97% | excl.^2^ |

***^a^****To test the effect of locus size in the QC, we grouped loci according to size (S; <0.6 Mb, M; 0.6-1.2 Mb, L; 1.2-1.8 Mb, XL; >3 Mb), keeping 15q11.2 separate given its high prevalence.* ***^b^****The number of probes used for rCNV calling from the Illumina PsychArray (PA) and Global Screening Array (GSA).* ***^c^****The number of genes overlapped >50% by the locus.* ***^d^****The sum of LOEUF scores of all locus genes (per-gene scores were inverted as lower LOEUF indicates increased gene constraint).* ***^e^****After processing raw PennCNV calls through our QCtreeCNV pipeline, all putative calls were verified by at least two analysts through visual inspection of probe intensities.* ***^f^****Each putative call was assigned as; true (T), false (F) or unknown (U), and we derived intra- and inter-rater reliability rates for (T) v. (F or U) assignments.* ***^g^****Loci with <5% rate of true calls (1), <5 total carriers from PA or GSA (2), or for which proportionality of hazards was not met in CPH models for the main iPSYCH2015 disorders (3), were excluded from further analysis.*

*Supplementary table 2: rCNV prevalence in iPSYCH2015 and the UKB*

|  | **Deletion prevalence** | | | | **Duplication prevalence** | | | |
| --- | --- | --- | --- | --- | --- | --- | --- | --- |
| **rCNV locus** | **iPSYCH (n)^a^** | **iPSYCH (%) [CI95%]^b^** | **UKB (%) [CI95%]^c^** | **P_FDR_^d^** | **iPSYCH (n)^a^** | **iPSYCH (%) [CI95%]^b^** | **UKB (%) [CI95%]^c^** | **P_FDR_^d^** |
| TAR | 25 | 0.029 [0.017-0.049] | 0.018 [0.016-0.020] | 0.18 | 175 | 0.109 [0.083-0.143] | 0.103 [0.098-0.108] | 0.74 |
| 1q21.1 | 66 | 0.032 [0.019-0.052] | 0.027 [0.024-0.029] | 0.62 | 136 | 0.066 [0.047-0.093] | 0.042 [0.039-0.045] | 0.081 |
| 2q11.2 | 17 | 0.006 [0.002-0.017] | 0.007 [0.006-0.009] | 0.71 | 6 | 0.009 [0.004-0.024] | 0.007 [0.006-0.008] | 0.72 |
| 2q13 | 57 | 0.049 [0.033-0.074] | 0.013 [0.011-0.015] | 1.0 × 10-6 | 25 | 0.020 [0.010-0.038] | 0.017 [0.015-0.019] | 0.74 |
| 2q21.1 | 18 | 0.010 [0.004-0.025] | 0.010 [0.008-0.011] | 0.90 | 14 | 0.019 [0.010-0.037] | 0.014 [0.012-0.016] | 0.62 |
| 10q11.23 | 23 | 0.024 [0.013-0.043] | 0.014 [0.012-0.015] | 0.14 | 13 | 0.010 [0.004-0.025] | 0.010 [0.009-0.012] | 0.95 |
| 13q12.12 | 17 | 0.012 [0.006-0.028] | 0.020 [0.018-0.023] | 0.36 | 23 | 0.013 [0.006-0.028] | 0.056 [0.053-0.060] | 0.0023 |
| 15q11.2 | 609 | 0.423 [0.368-0.485] | 0.395 [0.385-0.405] | 0.48 | 579 | 0.450 [0.393-0.515] | 0.484 [0.473-0.495] | 0.58 |
| PWAS | 5 | 0.009 [0.004-0.024] | na. | na. | 49 | 0.012 [0.006-0.026] | 0.005 [0.004-0.006] | 0.081 |
| 15q13.3 | 64 | 0.027 [0.016-0.046] | 0.010 [0.009-0.012] | 0.0037 | 104 | 0.070 [0.050-0.099] | 0.059 [0.056-0.063] | 0.58 |
| 16p13.11 | 129 | 0.079 [0.057-0.108] | 0.031 [0.029-0.034] | 2.1 × 10^-6^ | 294 | 0.174 [0.140-0.215] | 0.197 [0.190-0.204] | 0.58 |
| 16p12.1 | 163 | 0.122 [0.094-0.157] | 0.058 [0.055-0.062] | 2.1 × 10^-6^ | 47 | 0.044 [0.028-0.068] | 0.048 [0.045-0.052] | 0.74 |
| 16p11.2d | 48 | 0.037 [0.023-0.059] | 0.014 [0.012-0.016] | 7.5 × 10^-4^ | 68 | 0.045 [0.030-0.069] | 0.033 [0.030-0.035] | 0.51 |
| 16p11.2 | 81 | 0.048 [0.032-0.073] | 0.026 [0.024-0.029] | 0.013 | 185 | 0.103 [0.078-0.136] | 0.033 [0.030-0.036] | 1.1 × 10^-10^ |
| 17p12 | 45 | 0.048 [0.032-0.073] | 0.056 [0.053-0.060] | 0.59 | 24 | 0.024 [0.013-0.044] | 0.029 [0.027-0.032] | 0.72 |
| 17q12 | 28 | 0.016 [0.008-0.032] | 0.002 [0.002-0.003] | 1.9 × 10^-4^ | 65 | 0.032 [0.019-0.052] | 0.024 [0.022-0.027] | 0.58 |
| 22q11.2 | 36 | 0.018 [0.010-0.035] | 0.002 [0.002-0.003] | 3.3 × 10^-5^ | 156 | 0.083 [0.061-0.113] | 0.067 [0.063-0.071] | 0.54 |
| 22q11.2b | 23 | 0.013 [0.006-0.028] | na. | na. | 29 | 0.018 [0.009-0.035] | na. | na. |

***^a^****Total number of deletion and duplication carriers in iPSYCH2015 at the 18 herein studied rCNV loci.* ***^b^****Population-based prevalence and CI95% were calculated with svydesign() and svyciprop() functions from the survey package in R using finite population correction (fpc) to account for oversampling of cases.* ***^c^****rCNV prevalence in the UKB was calculated based on carrier counts from Crawford et al.^12^.* ***^d^****Differences in prevalence were assessed with a Welch’s test, and P-values adjusted for multiple comparisons with the false-discovery-rate (FDR) option of the p.adjust function in the stats package in R.*

*Supplementary table 3: rCNV-associated risk of psychiatric disorders in iPSYCH2015*

|  | **Deletion** | | **Duplication** | |  | **Deletion** | | **Duplication** | |
| --- | --- | --- | --- | --- | --- | --- | --- | --- | --- |
|  | **HR [CI95%]^a^** | **P** | **HR [CI95%]^a^** | **P** |  | **HR [CI95%]^a^** | **P** | **HR [CI95%]^a^** | **P** |
| **Locus** | **Attention-deficit hyperactivity disorder (ADHD)** | | | |  | **Autism spectrum disorder (ASD)** | | | |
| TAR | 0.59 [0.18-1.87] | 0.37 | 1.76 [1.15-2.69] | 0.0086 |  | 0.64 [0.2-2.03] | 0.44 | 1.49 [0.95-2.34] | 0.084 |
| 1q21.1 | 1.93 [0.92-4.07] | 0.083 | 2.65 [1.56-4.49] | 2.9E-04 |  | 2.37 [1.16-4.86] | 0.018 | 4.03 [2.44-6.64] | 4.6E-08 |
| 2q11.2 | 3.42 [0.88-13.2] | 0.075 | na.^b^ | na.^b^ |  | 2.44 [0.54-11] | 0.25 | na.^b^ | na.^b^ |
| 2q13 | 1.68 [0.83-3.36] | 0.15 | 1.43 [0.49-4.21] | 0.51 |  | 0.92 [0.4-2.08] | 0.84 | 0.97 [0.27-3.45] | 0.97 |
| 2q21.1 | 2.71 [0.69-10.6] | 0.15 | na.^b^ | na.^b^ |  | 2.49 [0.64-9.8] | 0.19 | 0.61 [0.12-3.14] | 0.55 |
| 10q11.23 | 0.32 [0.07-1.52] | 0.15 | na.^b^ | na.^b^ |  | 0.74 [0.22-2.51] | 0.63 | 1.81 [0.35-9.35] | 0.48 |
| 13q12.12 | 2.34 [0.82-6.68] | 0.11 | 2.65 [0.67-10.4] | 0.16 |  | 0.77 [0.15-3.98] | 0.75 | 5.00 [1.47-17.0] | 0.010 |
| 15q11.2 | 1.43 [1.14-1.8] | 0.0021 | 1.42 [1.13-1.79] | 0.0026 |  | 1.45 [1.15-1.84] | 0.0018 | 1.07 [0.82-1.38] | 0.63 |
| PWAS | na.^b^ | na.^b^ | 3.64 [1.04-12.8] | 0.044 |  | na.^b^ | na.^b^ | 20.8 [7.86-55.0] | 9.6E-10 |
| 15q13.3 | 2.51 [1.09-5.78] | 0.031 | 1.77 [0.98-3.19] | 0.057 |  | 3.74 [1.66-8.42] | 0.0014 | 1.59 [0.85-2.98] | 0.15 |
| 16p13.11 | 2.26 [1.37-3.73] | 0.0014 | 2.01 [1.43-2.81] | 5.0E-05 |  | 1.92 [1.12-3.31] | 0.018 | 1.84 [1.30-2.61] | 5.9E-04 |
| 16p12.1 | 1.86 [1.22-2.84] | 0.0039 | 0.56 [0.24-1.33] | 0.19 |  | 1.11 [0.68-1.83] | 0.68 | 1.00 [0.46-2.19] | 1.00 |
| 16p11.2d | 1.26 [0.55-2.88] | 0.58 | 1.57 [0.78-3.18] | 0.21 |  | 0.94 [0.37-2.39] | 0.89 | 1.26 [0.57-2.79] | 0.57 |
| 16p11.2 | 0.77 [0.36-1.61] | 0.48 | 2.63 [1.75-3.95] | 3.3E-06 |  | 3.07 [1.75-5.36] | 8.4E-05 | 2.04 [1.33-3.15] | 0.0012 |
| 17p12 | 0.45 [0.18-1.15] | 0.095 | 0.69 [0.21-2.23] | 0.53 |  | 0.81 [0.36-1.82] | 0.61 | 0.64 [0.17-2.41] | 0.51 |
| 17q12 | 4.24 [1.29-13.9] | 0.017 | 4.19 [1.98-8.87] | 1.8E-04 |  | 7.79 [2.71-22.4] | 1.4E-04 | 3.12 [1.37-7.08] | 0.0065 |
| 22q11.2 | 1.17 [0.49-2.8] | 0.72 | 2.61 [1.67-4.09] | 2.6E-05 |  | 1.5 [0.59-3.84] | 0.39 | 2.63 [1.66-4.16] | 3.4E-05 |
| 22q11.2b | 1.81 [0.53-6.18] | 0.35 | 1.63 [0.55-4.83] | 0.38 |  | 2.34 [0.67-8.13] | 0.18 | 0.92 [0.25-3.30] | 0.89 |
| **Locus** | **Major depressive disorder (MDD)** | | | |  | **Schizophrenia spectrum disorder (SSD)** | | | |
| TAR | 0.96 [0.28-3.28] | 0.95 | 1.45 [0.92-2.31] | 0.11 |  | 1.63 [0.45-5.83] | 0.45 | 1.02 [0.56-1.86] | 0.95 |
| 1q21.1 | 2.08 [0.86-5.04] | 0.10 | 1.43 [0.74-2.75] | 0.28 |  | 1.78 [0.68-4.69] | 0.24 | 2.7 [1.37-5.33] | 0.0042 |
| 2q11.2 | 2.18 [0.61-7.86] | 0.23 | na.^b^ | na.^b^ |  | 4.75 [1.07-21.1] | 0.041 | na.^b^ | na.^b^ |
| 2q13 | 1.39 [0.6-3.22] | 0.44 | 0.79 [0.2-3.04] | 0.73 |  | 1.58 [0.59-4.19] | 0.36 | 1.23 [0.29-5.18] | 0.78 |
| 2q21.1 | na.^b^ | na.^b^ | 0.52 [0.12-2.29] | 0.39 |  | 2.5 [0.28-22.8] | 0.42 | na.^b^ | na.^b^ |
| 10q11.23 | 0.77 [0.22-2.68] | 0.69 | 1 [0.18-5.66] | 1.00 |  | 0.66 [0.13-3.3] | 0.61 | 3.22 [0.6-17.1] | 0.17 |
| 13q12.12 | 0.39 [0.11-1.42] | 0.15 | 2.67 [0.56-12.8] | 0.22 |  | na.^b^ | na.^b^ | na.^b^ | na.^b^ |
| 15q11.2 | 1.17 [0.91-1.52] | 0.22 | 1.07 [0.82-1.38] | 0.62 |  | 0.99 [0.71-1.36] | 0.93 | 1.32 [0.97-1.78] | 0.077 |
| PWAS | na.^b^ | na.^b^ | 1.84 [0.31-10.9] | 0.50 |  | na.^b^ | na.^b^ | 9.19 [1.72-49.2] | 0.010 |
| 15q13.3 | 2.16 [0.84-5.5] | 0.11 | 1.54 [0.86-2.78] | 0.15 |  | 3.49 [1.36-8.95] | 0.0093 | 1.44 [0.71-2.95] | 0.31 |
| 16p13.11 | 1.06 [0.56-2.03] | 0.85 | 1.38 [0.94-2.01] | 0.10 |  | 2.29 [1.22-4.3] | 0.010 | 1.98 [1.31-3] | 0.0012 |
| 16p12.1 | 0.91 [0.54-1.52] | 0.71 | 0.86 [0.4-1.87] | 0.71 |  | 1.21 [0.66-2.2] | 0.54 | 0.48 [0.14-1.68] | 0.25 |
| 16p11.2d | 1.04 [0.4-2.73] | 0.94 | 1.17 [0.56-2.42] | 0.67 |  | 2.41 [0.96-6.08] | 0.062 | 1.81 [0.81-4.04] | 0.15 |
| 16p11.2 | 0.42 [0.18-1] | 0.049 | 1.04 [0.63-1.74] | 0.87 |  | 1.26 [0.55-2.88] | 0.58 | 1.5 [0.89-2.54] | 0.13 |
| 17p12 | 0.85 [0.38-1.91] | 0.70 | 0.55 [0.18-1.71] | 0.30 |  | 0.46 [0.13-1.65] | 0.24 | 1.17 [0.34-3.99] | 0.80 |
| 17q12 | na.^b^ | na.^b^ | 1.47 [0.53-4.07] | 0.46 |  | 4.84 [0.81-28.9] | 0.084 | 3.19 [1.17-8.68] | 0.023 |
| 22q11.2 | 1.03 [0.3-3.52] | 0.96 | 1.02 [0.59-1.77] | 0.94 |  | 3.28 [1.15-9.3] | 0.026 | 1.34 [0.7-2.54] | 0.37 |
| 22q11.2b | 1.11 [0.25-4.81] | 0.89 | 1.32 [0.43-4.08] | 0.63 |  | 2.6 [0.65-10.4] | 0.18 | 2.51 [0.77-8.16] | 0.12 |
| Table continues on next page | | | |  |  |  |  |  |  |
|  | **Deletion** | | **Duplication** | |  | **Deletion** | | **Duplication** | |
|  | **HR [CI95%]^a^** | **P** | **HR [CI95%]^a^** | **P** |  | **HR [CI95%]^a^** | **P** | **HR [CI95%]^a^** | **P** |
| **Locus** | **Any iPSYCH2015 disorder (ANY)** | | | |  | **Any affective disorder (AFF)** | | | |
| TAR | 0.68 [0.29-1.6] | 0.38 | 1.48 [1.02-2.14] | 0.037 |  | 0.9 [0.26-3.06] | 0.86 | 1.37 [0.87-2.18] | 0.18 |
| 1q21.1 | 2.12 [1.08-4.14] | 0.028 | 2.54 [1.58-4.08] | 0.0001 |  | 2.13 [0.89-5.09] | 0.087 | 1.45 [0.76-2.77] | 0.25 |
| 2q11.2 | 2.76 [0.93-8.12] | 0.067 | 0.31 [0.05-1.9] | 0.21 |  | 2.06 [0.57-7.43] | 0.27 | na.^b^ | na.^b^ |
| 2q13 | 1.28 [0.69-2.38] | 0.43 | 1.1 [0.43-2.82] | 0.85 |  | 1.29 [0.56-2.99] | 0.56 | 0.73 [0.19-2.82] | 0.65 |
| 2q21.1 | 2.34 [0.61-8.87] | 0.21 | 0.45 [0.14-1.45] | 0.18 |  | 1.51 [0.17-13.2] | 0.71 | 0.65 [0.17-2.53] | 0.53 |
| 10q11.23 | 0.71 [0.28-1.78] | 0.47 | 1.63 [0.44-6.05] | 0.46 |  | 0.72 [0.21-2.48] | 0.60 | 1.4 [0.3-6.57] | 0.67 |
| 13q12.12 | 0.71 [0.26-1.91] | 0.50 | 3.81 [1.25-11.6] | 0.019 |  | 0.36 [0.1-1.32] | 0.12 | 3.03 [0.66-13.9] | 0.15 |
| 15q11.2 | 1.3 [1.07-1.58] | 0.0085 | 1.21 [1-1.47] | 0.053 |  | 1.15 [0.89-1.49] | 0.28 | 1.09 [0.84-1.4] | 0.51 |
| PWAS | na.^b^ | na.^b^ | 11.3 [4.24-30.1] | 1.3E-06 |  | na.^b^ | na.^b^ | 1.7 [0.29-10.1] | 0.56 |
| 15q13.3 | 2.7 [1.31-5.56] | 0.0070 | 1.53 [0.94-2.5] | 0.091 |  | 2.01 [0.79-5.13] | 0.14 | 1.48 [0.82-2.66] | 0.19 |
| 16p13.11 | 1.75 [1.11-2.76] | 0.017 | 1.7 [1.27-2.27] | 0.0003 |  | 1.14 [0.61-2.14] | 0.69 | 1.42 [0.97-2.06] | 0.069 |
| 16p12.1 | 1.26 [0.86-1.84] | 0.24 | 0.81 [0.45-1.44] | 0.47 |  | 0.99 [0.6-1.62] | 0.96 | 0.87 [0.41-1.85] | 0.71 |
| 16p11.2d | 1.23 [0.62-2.44] | 0.56 | 1.32 [0.73-2.4] | 0.36 |  | 0.97 [0.37-2.54] | 0.95 | 1.2 [0.58-2.45] | 0.63 |
| 16p11.2 | na.^b^ | na.^b^ | na.^b^ | na.^b^ |  | 0.45 [0.2-1.03] | 0.057 | 0.99 [0.6-1.65] | 0.98 |
| 17p12 | 0.61 [0.33-1.15] | 0.13 | 0.64 [0.27-1.51] | 0.31 |  | 0.79 [0.35-1.77] | 0.57 | 0.52 [0.17-1.59] | 0.25 |
| 17q12 | na.^b^ | na.^b^ | na.^b^ | na.^b^ |  | 0.63 [0.08-5.11] | 0.68 | 1.56 [0.57-4.26] | 0.38 |
| 22q11.2 | 1.85 [0.83-4.14] | 0.13 | 1.85 [1.22-2.8] | 0.0035 |  | 0.95 [0.28-3.24] | 0.93 | 0.98 [0.57-1.7] | 0.95 |
| 22q11.2b | 1.64 [0.56-4.82] | 0.37 | 1.58 [0.64-3.89] | 0.32 |  | 1.03 [0.24-4.49] | 0.97 | 1.84 [0.66-5.16] | 0.25 |
| **Locus** | **Bipolar disorder (BPD)** | | | |  | **Schizophrenia (SCZ)** | | | |
| TAR | na.^b^ | na.^b^ | na.^b^ | na.^b^ |  | na.^b^ | na.^b^ | 1.5 [0.77-2.93] | 0.23 |
| 1q21.1 | 3.52 [0.88-14.1] | 0.076 | 1.63 [0.37-7.32] | 0.52 |  | 1.75 [0.52-5.84] | 0.36 | 2.36 [0.97-5.71] | 0.058 |
| 2q11.2 | na.^b^ | na.^b^ | na.^b^ | na.^b^ |  | 7.77 [1.25-48.2] | 0.028 | na.^b^ | na.^b^ |
| 2q13 | na.^b^ | na.^b^ | na.^b^ | na.^b^ |  | 1.39 [0.37-5.18] | 0.63 | 1.63 [0.31-8.59] | 0.57 |
| 2q21.1 | na.^b^ | na.^b^ | na.^b^ | na.^b^ |  | na.^b^ | na.^b^ | na.^b^ | na.^b^ |
| 10q11.23 | na.^b^ | na.^b^ | na.^b^ | na.^b^ |  | 1.3 [0.25-6.66] | 0.76 | na.^b^ | na.^b^ |
| 13q12.12 | na.^b^ | na.^b^ | na.^b^ | na.^b^ |  | na.^b^ | na.^b^ | 12.7 [2.17-74.1] | 0.0048 |
| 15q11.2 | 0.63 [0.3-1.31] | 0.22 | 1.26 [0.72-2.20] | 0.41 |  | 1.29 [0.88-1.88] | 0.19 | 1.25 [0.84-1.87] | 0.27 |
| PWAS | na.^b^ | na.^b^ | na.^b^ | na.^b^ |  | na.^b^ | na.^b^ | 6.88 [0.95-49.8] | 0.056 |
| 15q13.3 | na.^b^ | na.^b^ | 0.85 [0.19-3.79] | 0.84 |  | 3.28 [1.02-10.6] | 0.047 | 1.3 [0.5-3.37] | 0.59 |
| 16p13.11 | 2.83 [1.11-7.23] | 0.029 | 0.80 [0.28-2.27] | 0.67 |  | 1.77 [0.78-4.01] | 0.17 | 2.05 [1.22-3.45] | 0.0069 |
| 16p12.1 | 1.62 [0.65-4.03] | 0.30 | na.^b^ | na.^b^ |  | 1.34 [0.64-2.8] | 0.44 | 0.65 [0.15-2.87] | 0.56 |
| 16p11.2d | na.^b^ | na.^b^ | 1.89 [0.51-6.93] | 0.34 |  | 2.42 [0.78-7.52] | 0.13 | 2.38 [0.95-5.91] | 0.063 |
| 16p11.2 | na.^b^ | na.^b^ | 0.53 [0.13-2.28] | 0.40 |  | 0.57 [0.13-2.5] | 0.45 | 1.22 [0.59-2.52] | 0.58 |
| 17p12 | na.^b^ | na.^b^ | na.^b^ | na.^b^ |  | na.^b^ | na.^b^ | 2.38 [0.69-8.2] | 0.17 |
| 17q12 | na.^b^ | na.^b^ | 2.15 [0.39-11.8] | 0.38 |  | 5.38 [0.61-47.2] | 0.13 | 3.25 [0.98-10.7] | 0.054 |
| 22q11.2 | na.^b^ | na.^b^ | na.^b^ | na.^b^ |  | 3.18 [0.92-10.9] | 0.067 | 2.02 [0.99-4.12] | 0.053 |
| 22q11.2b | na.^b^ | na.^b^ | na.^b^ | na.^b^ |  | na.^b^ | na.^b^ | 2.56 [0.59-11] | 0.21 |

***^a^****Population-valid rCNV-associated hazard ratios (HR) and 95% confidence intervals (CI95%) were derived for each psychiatric disorder targeted in the iPSYCH2015 case-cohort design, using a Cox proportional hazards (CPH) model with inverse probability of sampling weights.* ***^b^****Comparisons involving <2 case carriers, or the CPH model failing a test of proportionality of hazards, were excluded.*

*Supplementary table 4: Comparison of rCNV-associated HRs of psychiatric disorders in iPSYCH2015 with Odds Ratios (ORs) reported in case-control studies*

| **rCNV** | **HR [CI95%]^a^** | **OR [CI95%]^b^** | **P_FDR_^c^** |  | **rCNV** | **HR [CI95%]^a^** | **OR [CI95%]^b^** | **P_FDR_^c^** |
| --- | --- | --- | --- | --- | --- | --- | --- | --- |
| **ADHD (case-control study; Gudmundsson *et al*.^4^)** | | | |  | **ASD (case-control study; Malhotra *et al*.^1^)** | | | |
| 15q11.2_del | 1.43 [1.14-1.80] | 1.65 [1.11-2.37] | 0.72 |  | 15q11.2_del | 1.45 [1.15-1.84] | 0.30 [0.10-1.40] | 0.081 |
| 15q13.3_del | 2.51 [1.09-5.78] | 5.97[ 2.63-12.6] | 0.46 |  | 15q13.3_del | 3.74 [1.66-8.42] | 10.8 [3.50-33.1] | 0.29 |
| 16p11.2_del | 0.77 [0.36-1.61] | 2.16 [0.75-5.11] | 0.41 |  | 16p11.2_del | 3.06 [1.75-5.36] | 9.50 [5.20-17.4] | 0.039 |
| 16p11.2_dup | 2.63 [1.75-3.95] | 4.34 [2.27-7.81] | 0.47 |  | 16p11.2_dup | 2.04 [1.33-3.15] | 11.8 [6.10-22.7] | 0.00014 |
| 16p11.2d_del | 1.26 [0.56-2.88] | 2.19[ 0.42-7.29] | 0.72 |  | 16p13.11_dup | 1.84 [1.30-2.61] | 1.50 [0.50-4.00] | 0.72 |
| 16p12.1_del | 1.86 [1.22-2.85] | 1.52 [0.63-3.16] | 0.72 |  | 17p12_del | 0.81 [0.36-1.82] | 4.00 [0.90-17.5] | 0.18 |
| 16p13.11_dup | 2.00 [1.43-2.81] | 2.12 [1.31-3.27] | 0.85 |  | 17q12_del | 7.79 [2.71-22.4] | 16.0 [2.90-87.9] | 0.66 |
| 17p12_del | 0.45 [0.18-1.15] | 2.20 [0.67-5.66] | 0.19 |  | 1q21.1_del | 2.37 [1.16-4.86] | 1.60 [0.20-11.7] | 0.72 |
| 17q12_dup | 4.19 [1.98-8.87] | 2.20 [0.76-5.24] | 0.65 |  | 1q21.1_dup | 4.03 [2.44-6.64] | 8.00 [3.50-18.4] | 0.31 |
| 1q21.1_del | 1.93 [0.92-4.07] | 2.68 [0.92-6.44] | 0.72 |  | 22q11.2_dup | 2.63 [1.67-4.16] | 3.30 [1.60-6.60] | 0.72 |
| 1q21.1_dup | 2.65 [1.56-4.49] | 3.44 [1.67-6.52] | 0.72 |  | PWAS_dup | 20.8 [7.86-55.0] | 42.6 [15.7-115] | 0.49 |
| 22q11.2_del | 1.17 [0.49-2.80] | 10.7 [4.66-23.2] | 0.0032 |  |  | | | |
| 22q11.2_dup | 2.61 [1.67-4.09] | 2.24 [1.32-3.63] | 0.72 |  | **SSD (case-control studies; Marshall *et al*.^5^, Rees *et al*.^6&7^)** | | | |
|  |  |  |  |  | 15q11.2_del^7^ | 0.98 [0.71-1.36] | 2.15 [1.71-2.68] | 0.0019 |
| **MDD (case-control study; Kendall *et al*.^10^)** | | | |  | 15q13.3_del^7^ | 3.49 [1.36-8.95] | 7.52 [3.98-14.2] | 0.44 |
| 15q11.2_del | 1.17 [0.91-1.52] | 1.11 [0.90-1.35] | 0.87 |  | PWAS_dup^7^ | 9.19 [1.72-49.2] | 13.2 [3.72-46.8] | 0.78 |
| 15q13.3_del | 2.15 [0.84-5.50] | 0.77 [0.13-2.52] | 0.56 |  | 16p11.2d_del^7^ | 2.41 [0.96-6.08] | 3.39 [1.21-9.52] | 0.78 |
| 16p11.2_del | 0.42 [0.18-1.00] | 1.2 1[0.54-2.34] | 0.49 |  | 16p13.11_dup^7^ | 1.98 [1.31-3.00] | 2.30 [1.57-3.36] | 0.78 |
| 16p11.2_dup | 1.04[ 0.63-1.74] | 2.65 [1.53-4.31] | 0.22 |  | 17p12_del^7^ | 0.46 [0.13-1.65] | 3.62 [1.73-7.57] | 0.029 |
| 16p11.2d_del | 1.04 [0.40-2.73] | 2.23 [0.92-4.63] | 0.56 |  | 17q12_del^7^ | 4.84 [0.81-28.8] | 6.64 [1.78-24.7] | 0.78 |
| 16p11.2d_dup | 1.17 [0.56-2.42] | 1.57 [0.82-2.73] | 0.79 |  | 1q21.1_del^7^ | 1.78 [0.68-4.69] | 8.35 [4.65-15.0] | 0.029 |
| 16p12.1_del | 0.91 [0.54-1.52] | 1.47 [0.90-2.27] | 0.49 |  | 1q21.1_dup^7^ | 2.70 [1.37-5.33] | 3.45 [1.92-6.20] | 0.78 |
| 16p13.11_del | 1.06 [0.56-2.02] | 2.21 [1.25-3.63] | 0.49 |  | TAR_del^6^ | 1.63[ 0.45-5.83] | 1.20 [0.36-3.90] | 0.78 |
| 16p13.11_dup | 1.38 [0.94-2.01] | 0.87 [0.63-1.78] | 0.49 |  | TAR_dup^6^ | 1.02 [0.56-1.86] | 1.90 [0.93-3.93] | 0.44 |
| 17q12_dup | 1.47 [0.53-4.07] | 1.55 [0.69-3.02] | 0.98 |  | 2q11.2_del^6^ | 4.75 [1.07-21.1] | 9.30 [1.03-447] | 0.78 |
| 1q21.1_del | 2.08 [0.86-5.04] | 1.11 [0.46-2.22] | 0.56 |  | 2q13_del^6^ | 1.58 [0.59-4.18] | 3.60 [0.26-206] | 0.78 |
| 1q21.1_dup | 1.43 [0.74-2.75] | 2.17 [1.34-3.36] | 0.56 |  | 2q13_dup^6^ | 1.23 [0.29-5.18] | 1.70 [0.30-9.98] | 0.78 |
| 22q11.2_del | 1.03 [0.30-3.52] | 1.69 [0.09-9.17] | 0.87 |  | 16p11.2d_dup^6^ | 1.81 [0.81-4.04] | 1.20 [0.44-3.08] | 0.78 |
| 22q11.2_dup | 1.02 [0.59-1.77] | 1.72 [1.12-2.53] | 0.49 |  | 16p11.2_del^6^ | 1.26 [0.55-2.88] | 0.62 [0.19-1.79] | 0.64 |
| 2q11.2_del | 2.18[ 0.60-7.86] | 2.34 [0.69-6.02] | 0.98 |  | 16p12.1_del^6^ | 1.21 [0.66-2.20] | 3.30 [1.61-7.05] | 0.12 |
| 2q13_del | 1.39 [0.60-3.22] | 0.98 [0.24-2.67] | 0.85 |  | 16p13.11_del^6^ | 2.29 [1.22-4.30] | 1.10 [0.43-2.73] | 0.44 |
| 2q13_dup | 0.79 [0.20-3.04] | 1.29 [0.49-2.90] | 0.79 |  | 17q12_dup^6^ | 3.19 [1.17-8.68] | 2.20 [0.74-6.76] | 0.78 |
| PWAS_dup | 1.84 [0.31-10.9] | 8.14 [2.77-21.7] | 0.49 |  | 16p11.2_dup^5^ | 1.50 [0.89-2.54] | 9.40 [4.20-20.9] | 0.0019 |
| TAR_del | 0.96 [0.28-3.28] | 0.95 [0.29-2.29] | 0.99 |  | 22q11.2_del^5^ | 3.28 [1.15-9.30] | 67.7 [9.40-492] | 0.029 |
| TAR_dup | 1.45 [0.92-2.31] | 1.17 [0.77-1.69] | 0.79 |  | 22q11.2_dup^5^ | 1.34 [0.70-2.54] | 0.15 [0.04-0.52] | 0.021 |

*^a^rCNV-associated HRs and 95% confidence intervals (CI95%) for ASD, ADHD, MDD, and SSD in iPSYCH2015 case-cohort are shown in the table, in case they had comparable risk estimates reported in case-control studies. ^b^ Odds ratios (ORs) and 95% confidence intervals (CI95%) of rCNV-associated risk of ASD, ADHD, MDD, and SSD reported in published case-control studies. ^C^Difference between HRs in iPSYCH2015 and previously reported ORs was evaluated with a Welch’s t-test and P-values adjusted for multiple comparisons for each disorder separately with a false discovery rate (FDR) test using the p.adjust() function from the stat package in R.*

*Supplementary table 5: Comparison of Hazard ratios (HRs) with Odds ratios (ORs) associated with rCNVs across all the studied outcomes in iPSYCH2015 case-cohort*

See: [Supplementary_table_5.xlsx](https://docs.google.com/spreadsheets/d/1f5DgDPSXxPbYXHQxyk-602u85rrEWbnP/edit?usp=sharing&ouid=116231106222144114965&rtpof=true&sd=true)

*Supplementary table 6 Diagnosis-specific rCNV effects on risk between iPSYCH2015 disorders*

| **Locus** | **rCNV** | | **ASD v. ADHD** ^a^ | **P_FDR_ ^b^** | **SSD v. ADHD ^a^** | **P_FDR_ ^b^** | **SSD v. ASD ^a^** | **P_FDR_ ^b^** |
| --- | --- | --- | --- | --- | --- | --- | --- | --- |
| TAR | del | | -0.27 | 0.8363 | 1.28 | 0.3115 | 1.55 | 0.0324 |
|  | dup | | -0.35 | 0.3902 | -0.79 | 0.120 | -0.44 | 0.4877 |
| 1q21.1 | del | | 0.24 | 0.7626 | -0.39 | 0.7065 | -0.65 | 0.5233 |
|  | dup | | 0.75 | 0.0821 | -0.32 | 0.6001 | -1.04 | 0.0330 |
| 2q11.2 | del | | -0.60 | 0.7226 | 0.53 | 0.7077 | 1.12 | 0.5822 |
|  | dup | | na.^c^ | na.^c^ | na.^c^ | na.^c^ | na.^c^ | na.^c^ |
| 2q13 | del | | -0.69 | 0.2962 | -0.29 | 0.7077 | 0.35 | 0.7802 |
|  | dup | | -0.59 | 0.7210 | -0.31 | 0.7802 | 0.29 | 0.9148 |
| 2q21.1 | del | | -0.03 | 0.9753 | -1.08 | 0.4956 | -0.95 | 0.6774 |
|  | dup | | na.^c^ | na.^c^ | na.^c^ | na.^c^ | na.^c^ | na.^c^ |
| 10q11.23 | del | | 1.90 | 0.2962 | 1.69 | 0.4383 | -0.18 | 0.9549 |
|  | dup | | na.^c^ | na.^c^ | na.^c^ | na.^c^ | 0.68 | 0.7802 |
| 13q12.12 | del | | -1.98 | 0.0593 | na.^c^ | na.^c^ | na.^c^ | na.^c^ |
|  | dup | | 1.14 | 0.3486 | na. | na.^c^ | na.^c^ | na.^c^ |
| 15q11.2 | del | | 0.02 | 0.9021 | -0.53 | 0.0327 | -0.56 | 0.0252 |
|  | dup | | -0.43 | 0.0495 | -0.17 | 0.5873 | 0.25 | 0.4559 |
| PWAS | del | | na.^c^ | na.^c^ | na.^c^ | na.^c^ | na.^c^ | na.^c^ |
|  | dup | | 3.53 | 2.2×10^-16^ | 0.71 | 0.5836 | -2.45 | 0.0013 |
| 15q13.3 | del | | 0.74 | 0.3077 | 0.31 | 0.7077 | -0.39 | 0.7595 |
|  | dup | | -0.24 | 0.7226 | -0.08 | 0.8764 | 0.15 | 0.8990 |
| 16p13.11 | del | | -0.40 | 0.4148 | -0.13 | 0.7936 | 0.24 | 0.7786 |
|  | dup | | -0.13 | 0.7226 | -0.13 | 0.7077 | 0.00 | 0.9979 |
| 16p12.1 | del | | -0.83 | 0.0445 | -0.61 | 0.2321 | 0.19 | 0.8124 |
|  | dup | | 0.92 | 0.3326 | 0.09 | 0.9135 | -0.81 | 0.4877 |
| 16p11.2d | del | | -0.28 | 0.780 | 0.91 | 0.3867 | 1.18 | 0.2432 |
|  | dup | | -0.51 | 0.5194 | 0.31 | 0.7077 | 0.80 | 0.4244 |
| 16p11.2 | del | | 2.17 | 0.0001 | 0.64 | 0.4956 | -1.42 | 0.0282 |
|  | dup | | -0.57 | 0.1171 | -0.88 | 0.0327 | -0.38 | 0.4973 |
| 17p12 | del | | 0.78 | 0.3077 | -0.35 | 0.7802 | -1.11 | 0.4544 |
|  | dup | | -0.14 | 0.9021 | 0.91 | 0.5524 | 1.05 | 0.4544 |
| 17q12 | del | | 1.10 | 0.2860 | -0.15 | 0.8764 | -1.11 | 0.4345 |
|  | dup | | -0.65 | 0.3411 | -0.64 | 0.4882 | -0.05 | 0.9549 |
| 22q11.2 | del | | 0.43 | 0.7226 | 1.26 | 0.2321 | 0.84 | 0.4639 |
|  | dup | | -0.14 | 0.780 | -1.05 | 0.0227 | -0.91 | 0.0788 |
| 22q11.2b | del | | 0.24 | 0.8377 | 0.63 | 0.6731 | 0.39 | 0.8124 |
|  | dup | | -0.83 | 0.4999 | 0.47 | 0.7077 | 1.26 | 0.4244 |

*^a^Coefficients of the interaction term between diagnosis and rCNV were extracted from the results of GEE models corresponding to three pairwise comparisons across ASD, ADHD, and SSD using glmgee() function from the glmtoolbox package in R (see Method). ^b^P-values corresponding to each coefficient of the interaction term between diagnosis and rCNV within each pairwise diagnosis were retrieved from the GEE models and adjusted for multiple comparisons with the false-discovery-rate (FDR) by p.adjust() function from the stat package in R. ^c^ We included only those rCNVs in each GEE analysis that had valid HR estimates for the two compared diagnoses within each pairwise comparison.*

*Supplementary table 7: rCNV-associated risk of ID and epilepsy in iPSYCH2015*

|  | **Deletion** | | **Duplication** | |  | **Deletion** | | **Duplication** | |
| --- | --- | --- | --- | --- | --- | --- | --- | --- | --- |
|  | **HR [CI95%]^a^** | **P** | **HR [CI95%]^a^** | **P** |  | **HR [CI95%]^a^** | **P** | **HR [CI95%]^a^** | **P** |
| **Locus** | **Intellectual disability (ID)** | | | |  | **Epilepsy (EPI)** | | | |
| TAR | 1.18 [0.26-5.29] | 0.83 | 1.18 [0.55-2.51] | 0.67 |  | na.^b^ | na.^b^ | 1.49 [0.65-3.42] | 0.34 |
| 1q21.1 | 5.38 [2.40-12.1] | 4.6 × 10^-5^ | 3.14 [1.59-6.20] | 9.9 × 10^-4^ |  | 1.63 [0.41-6.47] | 0.49 | 1.79 [0.66-4.88] | 0.25 |
| 2q11.2 | 13.4 [2.97-60.7] | 7.4 × 10^-4^ | na.^b^ | na.^b^ |  | 9.52 [2.42-37.4] | 0.0013 | na.^b^ | na.^b^ |
| 2q13 | 1.54 [0.52-4.56] | 0.44 | 0.83 [0.10-6.68] | 0.86 |  | 0.64 [0.09-4.69] | 0.66 | na.^b^ | na.^b^ |
| 2q21.1 | 4.67 [1.27-17.2] | 0.021 | 2.05 [0.41-10.1] | 0.38 |  | 4.17 [0.48-36.1] | 0.20 | na.^b^ | na.^b^ |
| 10q11.23 | 0.67 [0.08-5.29] | 0.70 | 1.97 [0.22-17.72] | 0.54 |  | na.^b^ | na.^b^ | 3.15 [0.36-27.8] | 0.30 |
| 13q12.12 | 2.40 [0.48-12.1] | 0.29 | 2.09 [0.25-17.3] | 0.49 |  | 3.20 [0.65-15.8] | 0.15 | 3.24 [0.59-17.7] | 0.17 |
| 15q11.2 | 1.46 [1.02-2.07] | 0.038 | 1.25 [0.86-1.83] | 0.24 |  | 1.90 [1.29-2.79] | 0.0010 | 0.91 [0.53-1.55] | 0.72 |
| PWAS | na.^b^ | na.^b^ | 46.5 [20.1-108] | 3.5 × 10^-19^ |  | na.^b^ | na.^b^ | 15.1 [7.38-30.9] | 1.1 × 10^-13^ |
| 15q13.3 | 10.4 [5.03-21.5] | 2.7 × 10^-10^ | 2.28 [1.04-5.00] | 0.041 |  | 7.33 [3.32-16.2] | 8.1 × 10^-7^ | 1.70 [0.62-4.62] | 0.30 |
| 16p13.11 | 6.12 [3.63-10.3] | 1.1 × 10^-11^ | 2.04 [1.24-3.35] | 0.0051 |  | 10.0 [6.60-15.2] | 3.2 × 10^-27^ | 0.50 [0.16-1.56] | 0.23 |
| 16p12.1 | 1.49 [0.75-2.96] | 0.25 | 1.24 [0.36-4.26] | 0.73 |  | 1.64 [0.76-3.56] | 0.21 | 0.63 [0.09-4.65] | 0.65 |
| 16p11.2d | 2.76 [1.07-7.11] | 0.036 | 1.86 [0.69-5.01] | 0.22 |  | 5.26 [2.17-12.8] | 2.4 × 10^-4^ | 1.20 [0.28-5.07] | 0.81 |
| 16p11.2 | 5.14 [2.67-9.91] | 1.0 × 10^-6^ | 4.43 [2.76-7.09] | 5.9 × 10^-10^ |  | 5.47 [2.59-11.6] | 8.7 × 10^-6^ | 2.35 [1.20-4.62] | 0.013 |
| 17p12 | 0.33 [0.04-2.49] | 0.28 | 2.80 [0.88-8.86] | 0.080 |  | na.^b^ | na.^b^ | 3.35 [1.00-11.2] | 0.050 |
| 17q12 | 16.1 [5.90-43.8] | 5.6 × 10^-8^ | 2.96 [1.06-8.25] | 0.038 |  | na.^b^ | na.^b^ | 2.01 [0.48-8.45] | 0.34 |
| 22q11.2 | 18.5 [7.98-42.7] | 9.6 × 10^-12^ | 3.80 [2.15-6.71] | 4.4 × 10^-6^ |  | 11.4 [5.36-24.4] | 2.8 × 10^-10^ | 1.97 [0.85-4.60] | 0.12 |
| 22q11.2b | 1.66 [0.20-13.8] | 0.64 | na.^b^ | na.^b^ |  | 6.38 [2.03-20.1] | 0.0015 | 1.48 [0.20-11.2] | 0.70 |

***^a^****rCNV-associated hazard ratios (HR) and 95% confidence intervals (CI95%) were derived for intellectual disability and epilepsy using a sex-stratified Cox proportional hazards (CPH) model with inverse probability of sampling (IPS) weights.* ***^b^****Comparisons involving <2 case carriers, or the CPH model failing a test of proportionality of hazards, were excluded.*

*Supplementary table 8: Factors affecting initial evaluation of rCNV calls*

| **Predictor** | **Level** | **N (calls)** | **pU^a^** | **iIRR^a^** | **Anova model^b^** |
| --- | --- | --- | --- | --- | --- |
| LRR-SD | 0.10-0.15 | 350 | 0.003 | 0.014 | pU; deviance = 221.0, P = 5.5 × 10^-50^  iIRR; deviance = 210.6, P = 1.0 × 10^-47^ |
|  | 0.15-0.20 | 3,435 | 0.009 | 0.012 |  |
|  | 0.20-0.25 | 2,369 | 0.023 | 0.031 |  |
|  | 0.25-0.30 | 962 | 0.097 | 0.097 |  |
|  | 0.30-0.35 | 257 | 0.136 | 0.148 |  |
| Locus group | 15q11.2 | 1,253 | 0.015 | 0.019 | pU; deviance = 199.8, P = 9.5 × 10^-29^  iIRR; deviance = 205.0, P = 9.7 × 10^-30^ |
|  | S (<0.6 Mb) | 1,732 | 0.073 | 0.078 |  |
|  | M (0.6-1.2 Mb) | 1,318 | 0.041 | 0.054 |  |
|  | L (1.2-1.8 Mb) | 2,849 | 0.005 | 0.006 |  |
|  | XL (>3 Mb) | 221 | 0.014 | 0.009 |  |
| Genotyping array | PsychArray | 3,022 | 0.050 | 0.050 | pU; deviance = 24.7, P = 6.8 × 10^-7^  iIRR; deviance = 7.4, P = 0.0064 |
|  | GSA | 4,351 | 0.015 | 0.023 |  |
| CNV type | Deletion | 2,553 | 0.045 | 0.056 | pU; deviance = 0.1, P = 0.80  iIRR; deviance = 1.0, P = 0.31 |
|  | Duplication | 4,820 | 0.021 | 0.022 |  |

*All rCNV calls were visually inspected by at least two analysts and evaluated as; true (T), false (F) or unknown/undetermined (U). We derived inter-rater-reliability (IRR) by comparing consensus between raters (T v. F/U).* ***^a^****The fraction of calls rated as unknown (pU) and of calls without consensus between raters during initial rating (iIRR) for bins of increasing LRR-SD, and of each category of the other call properties used to predict pU and iIRR in the initial QC analysis.* ***^b^****Calls of undetermined status (U) and calls without consensus across raters during initial rating (iIRR) were predicted using different call properties; sample LRR-SD, Locus group, Genotyping array, and CNV type (deletion or duplication). Each inspected call was assigned a 0 or 1 for unknown status (U) and a 0 or 1 for lacking initial inter-rater consensus (iIRR). Predictor variables were added sequentially to a logistic model according to significance and significance re-evaluted by comparison with the corresponding nested model with anova.*

*Supplementary table 9: Prevalence of rCNVs across bins of increasing LRR-SD*

| **Variable^a^** | **Class^a^** | **LRR-SD^b^** | **N (carriers)^c^** | **N (samples)^d^** | **Prevalence (%) [CI95]^e^** | **P^f^** |
| --- | --- | --- | --- | --- | --- | --- |
| All rCNVs | All carriers | 0.10 - 0.15 | 297 | 10,687 | 2.53 [ 2.12 - 3.02 ] | 0.42 |
|  |  | 0.15 - 0.20 | 1,626 | 56,170 | 2.30 [ 2.11 - 2.50 ] |  |
|  |  | 0.20 - 0.25 | 1,132 | 37,563 | 2.38 [ 2.15 - 2.65 ] |  |
|  |  | 0.25 - 0.30 | 361 | 12,890 | 2.18 [ 1.81 - 2.63 ] |  |
|  |  | 0.30 - 0.35 | 78 | 2,937 | 1.84 [ 1.21 - 2.80 ] |  |
| rCNV type | Deletion carriers | 0.10 - 0.15 | 117 | 10,687 | 0.97 [ 0.73 - 1.29 ] | 0.69 |
|  |  | 0.15 - 0.20 | 677 | 56,170 | 1.03 [ 0.91 - 1.17 ] |  |
|  |  | 0.20 - 0.25 | 497 | 37,563 | 1.06 [ 0.91 - 1.24 ] |  |
|  |  | 0.25 - 0.30 | 163 | 12,890 | 0.95 [ 0.71 - 1.26 ] |  |
|  |  | 0.30 - 0.35 | 35 | 2,937 | 0.86 [ 0.46 - 1.60 ] |  |
|  | Duplication carriers | 0.10 - 0.15 | 183 | 10,687 | 1.57 [ 1.25 - 1.96 ] | 0.21 |
|  |  | 0.15 - 0.20 | 960 | 56,170 | 1.28 [ 1.14 - 1.43 ] |  |
|  |  | 0.20 - 0.25 | 653 | 37,563 | 1.36 [ 1.18 - 1.56 ] |  |
|  |  | 0.25 - 0.30 | 201 | 12,890 | 1.26 [ 0.98 - 1.61 ] |  |
|  |  | 0.30 - 0.35 | 43 | 2,937 | 0.98 [ 0.55 - 1.74 ] |  |
| Genotyping array | GSA  (all carriers) | 0.10 - 0.15 | 152 | 6,180 | 2.00 [ 1.52 - 2.63 ] | 0.84 |
|  |  | 0.15 - 0.20 | 870 | 30,472 | 2.31 [ 2.06 - 2.59 ] |  |
|  |  | 0.20 - 0.25 | 193 | 7,765 | 2.04 [ 1.60 - 2.61 ] |  |
|  |  | 0.25 - 0.30 | 38 | 1,537 | 1.88 [ 1.07 - 3.27 ] |  |
|  |  | 0.30 - 0.35 | 9 | 436 | 2.70 [ 1.06 - 6.68 ] |  |
|  | PsychArray  (all carriers) | 0.10 - 0.15 | 145 | 4,507 | 3.10 [ 2.46 - 3.90 ] | 0.16 |
|  |  | 0.15 - 0.20 | 756 | 25,698 | 2.29 [ 2.02 - 2.60 ] |  |
|  |  | 0.20 - 0.25 | 939 | 29,798 | 2.48 [ 2.21 - 2.78 ] |  |
|  |  | 0.25 - 0.30 | 323 | 11,353 | 2.23 [ 1.83 - 2.71 ] |  |
|  |  | 0.30 - 0.35 | 69 | 2,501 | 1.69 [ 1.05 - 2.70 ] |  |
| Locus group | Carriers of 15q11.2 CNVs | 0.10 - 0.15 | 99 | 10,687 | 0.98 [ 0.73 - 1.30 ] | 0.42 |
|  |  | 0.15 - 0.20 | 568 | 56,170 | 0.90 [ 0.78 - 1.03 ] |  |
|  |  | 0.20 - 0.25 | 375 | 37,563 | 0.87 [ 0.73 - 1.04 ] |  |
|  |  | 0.25 - 0.30 | 114 | 12,890 | 0.69 [ 0.50 - 0.97 ] |  |
|  |  | 0.30 - 0.35 | 32 | 2,937 | 0.76 [ 0.39 - 1.46 ] |  |
|  | Carriers of other small CNVs  (<0.6 Mb) | 0.10 - 0.15 | 77 | 10,687 | 0.59 [ 0.40 - 0.85 ] | 0.87 |
|  |  | 0.15 - 0.20 | 388 | 56,170 | 0.57 [ 0.48 - 0.68 ] |  |
|  |  | 0.20 - 0.25 | 246 | 37,563 | 0.54 [ 0.43 - 0.67 ] |  |
|  |  | 0.25 - 0.30 | 90 | 12,890 | 0.55 [ 0.37 - 0.79 ] |  |
|  |  | 0.30 - 0.35 | 20 | 2,937 | 0.72 [ 0.36 -1.44 ] |  |
|  | Carriers of medium sized CNVs (0.6-1.2 Mb) | 0.10 - 0.15 | 66 | 10,687 | 0.49 [ 0.33 - 0.73 ] | 0.92 |
|  |  | 0.15 - 0.20 | 352 | 56,170 | 0.44 [ 0.36 - 0.53 ] |  |
|  |  | 0.20 - 0.25 | 283 | 37,563 | 0.59 [ 0.47 - 0.72 ] |  |
|  |  | 0.25 - 0.30 | 83 | 12,890 | 0.41 [ 0.27 - 0.63 ] |  |
|  |  | 0.30 - 0.35 | 14 | 2,937 | 0.23 [ 0.07 - 0.73 ] |  |
|  | Carriers of large CNVs (>1.2 Mb) | 0.10 - 0.15 | 60 | 10,687 | 0.51 [ 0.34 - 0.76 ] | 0.78 |
|  |  | 0.15 - 0.20 | 345 | 56,170 | 0.41 [ 0.34 - 0.51 ] |  |
|  |  | 0.20 - 0.25 | 251 | 37,563 | 0.45 [ 0.35 - 0.57 ] |  |
|  |  | 0.25 - 0.30 | 77 | 12,890 | 0.56 [ 0.38 - 0.81 ] |  |
|  |  | 0.30 - 0.35 | 12 | 2,937 | 0.13 [ 0.03 - 0.56 ] |  |

***^a^****rCNVs carriers were classified by dosage type, genotyping array and locus size group.* ***^b^****Samples were binned by increasing LRR-SD, a main indicator of sample quality.* ***^c^****Number of rCNV carriers and* ***^d^****corresponding number of samples of each class in each LRR-SD bin.* ***^e^****The population-based rCNV prevalence and 95% confidence interval for each class for each LRR-SD bin, calculated using finite population correction weights with the svydesign() and svyciprop() functions from the survey package in R.* ***^f^****P-value from a logistic regression model predicting carrier status for each respective CNV class from per-sample LRR-SD (using the svyglm() function from the survey package in R).*

*Supplementary table 10: Prevalence of rCNVs by genotyping array*

| **Locus** | **PsychArray** | | **GSA** | | **P^c^** |
| --- | --- | --- | --- | --- | --- |
|  | **N (carriers)^a^** | **Prevalence (%) [CI95]^b^** | **N (carriers)^a^** | **Prevalence (%) [CI95]^b^** |  |
| All CNVs (all loci) | 2,232 | 2.40 [ 2.23 - 2.58 ] | 1,262 | 2.21 [ 2.01 - 2.43 ] | 0.18 |
| Deletions (all loci) | 957 | 1.07 [0.96 - 1.19 ] | 532 | 0.94 [ 0.82 - 1.09 ] | 0.18 |
| Duplications (all loci) | 1,304 | 1.35 [1.23 - 1.49 ] | 736 | 1.28 [ 1.13 - 1.45 ] | 0.48 |
| Small (<0.6 Mb) | 512 | 0.57 [ 0.49 - 0.66 ] | 309 | 0.56 [ 0.47 - 0.68 ] | 0.97 |
| Medium (0.6-1.2 Mb) | 518 | 0.52 [ 0.44 - 0.61 ] | 280 | 0.42 [ 0.34 - 0.52 ] | 0.13 |
| Large (>1.2 Mb) | 499 | 0.44 [ 0.37 - 0.52 ] | 246 | 0.44 [ 0.36 - 0.55 ] | 0.98 |
| TAR | 125 | 0.133 [ 0.097 - 0.182 ] | 98 | 0.145 [ 0.100 - 0.211 ] | 0.73 |
| 1q21.1 | 144 | 0.117 [ 0.084 - 0.163 ] | 58 | 0.067 [ 0.040 - 0.115 ] | 0.083 |
| 2q11.2 | 17 | 0.021 [ 0.009 - 0.046 ] | 6 | 0.007 [ 0.001 - 0.036 ] | 0.24 |
| 2q13 | 51 | 0.065 [ 0.042 - 0.103 ] | 31 | 0.074 [ 0.043 - 0.126 ] | 0.73 |
| 2q21.1 | 16 | 0.017 [ 0.007 - 0.040 ] | 17 | 0.049 [ 0.025 - 0.094 ] | 0.058 |
| 3q29 | 22 | 0.010 [ 0.003 - 0.028 ] | 7 | 0.007 [ 0.001 - 0.036 ] | 0.71 |
| 10q11.23 | 22 | 0.040 [ 0.022 - 0.072 ] | 14 | 0.025 [ 0.010 - 0.062 ] | 0.41 |
| 13q12.12 | 27 | 0.033 [ 0.017 - 0.062 ] | 13 | 0.014 [ 0.004 - 0.044 ] | 0.20 |
| 15q11.2 | 743 | 0.911 [ 0.808 - 1.030 ] | 445 | 0.814 [ 0.695 - 0.953 ] | 0.27 |
| PWAS | 43 | 0.027 [ 0.014 - 0.053 ] | 11 | 0.013 [ 0.004 - 0.044 ] | 0.31 |
| 15q13.1 | 13 | 0.013 [ 0.005 - 0.035 ] | 9 | 0.007 [ 0.002 - 0.035 ] | 0.56 |
| 15q13.3 | 107 | 0.087 [ 0.059 - 0.127 ] | 61 | 0.113 [ 0.074 - 0.173 ] | 0.36 |
| 16p13.11 | 264 | 0.265 [ 0.212 - 0.331 ] | 159 | 0.233 [ 0.174 - 0.312 ] | 0.50 |
| 16p12.1 | 134 | 0.164 [ 0.123 - 0.218 ] | 77 | 0.167 [ 0.118 - 0.238 ] | 0.93 |
| 16p11.2d | 77 | 0.087 [ 0.059 - 0.128 ] | 40 | 0.076 [ 0.045 - 0.127 ] | 0.67 |
| 16p11.2 | 169 | 0.168 [ 0.127 - 0.222 ] | 97 | 0.126 [ 0.085 - 0.186 ] | 0.23 |
| 17p12 | 48 | 0.072 [ 0.047 - 0.112 ] | 21 | 0.072 [ 0.042 - 0.124 ] | 0.99 |
| 17q12 | 58 | 0.044 [ 0.026 - 0.075 ] | 35 | 0.052 [ 0.028 - 0.096 ] | 0.69 |
| 22q11.2 | 124 | 0.081 [ 0.055 - 0.120 ] | 68 | 0.132 [ 0.089 - 0.196 ] | 0.087 |
| 22q11.2b | 31 | 0.033 [ 0.018 - 0.062 ] | 21 | 0.026 [ 0.011 - 0.062 ] | 0.67 |

***^a^****Total number of rCNV carriers of each class and locus genotyped on each array (out of a total of 73,857 and 46,390 individuals genotyped on the Illumina PsychArray and GSA, respectively).* ***^b^****Population-based CNV prevalence (with 95% CI) for each class, calculated using finite population correction weights with the svydesign() and svyciprop() functions from the survey package in R.* ***^c^****The last column details the P-value from a logistic regression model predicting carrier status for each respective CNV class from genotyping array (using the svyglm() function from the survey package in R.*
