## Supplementary table 5 for "Population-based Risk of Psychiatric Disorders Associated with Recurrent CNVs"

*Supplementary Table 5: Comparison of Hazard ratios (HRs) with Odds ratios (ORs) associated with rCNVs across all the studied outcomes in iPSYCH2015 case-cohort. For sensitivity analysis, we computed rCNV-associated ORs with 95% confidence intervals (CI95%) using generalised linear models (GLMs) to compare with weighted Cox proportional HRs (see Method). ORs are only reported in case of corresponding valid HRs in our study.*

| | | Deletion | | Duplication | | Deletion | | Duplication | |
|---|---|---|---|---|---|---|---|---|---|
| **Locus** | **Diagnosis** | **HR [CI95%]** | **P** | **HR [CI95%]** | **P** | **OR [CI95%]** | **P** | **OR [CI95%]** | **P** |
| TAR | ADHD | 0.59 [0.18-1.87] | 0.37 | 1.76 [1.15-2.69] | 0.0086 | 0.47 [0.13-1.39] | 0.20 | 1.85 [1.23-2.79] | 0.0034 |
| 1q21.1 | ADHD | 1.93 [0.92-4.07] | 0.083 | 2.65 [1.56-4.49] | 2.90E-04 | 1.99 [0.99-4.1] | 0.057 | 2.22 [1.37-3.64] | 0.0012 |
| 2q11.2 | ADHD | 3.42 [0.88-13.2] | 0.075 | na. | na. | 2.45 [0.62-10.3] | 0.20 | na. | na. |
| 2q13 | ADHD | 1.68 [0.83-3.36] | 0.15 | 1.43 [0.49-4.21] | 0.51 | 1.24 [0.65-2.37] | 0.51 | 1.18 [0.41-3.38] | 0.75 |
| 2q21.1 | ADHD | 2.71 [0.69-10.6] | 0.15 | na. | na. | 2.52 [0.75-9.75] | 0.15 | na. | na. |
| 10q11.23 | ADHD | 0.32 [0.07-1.52] | 0.15 | na. | na. | 0.31 [0.05-1.21] | 0.14 | na. | na. |
| 13q12.12 | ADHD | 2.34 [0.82-6.68] | 0.11 | 2.65 [0.67-10.44] | 0.16 | 3.63 [1.21-12.12] | 0.025 | 1.33 [0.32-5.21] | 0.68 |
| 15q11.2 | ADHD | 1.43 [1.14-1.8] | 0.0021 | 1.42 [1.13-1.79] | 0.0026 | 1.48 [1.18-1.84] | 0.0005 | 1.32 [1.06-1.64] | 0.015 |
| PWAS | ADHD | na. | na. | 3.64 [1.04-12.8] | 0.044 | na. | na. | 3.06 [1.03-10.19] | 0.051 |
| 15q13.3 | ADHD | 2.51 [1.09-5.78] | 0.031 | 1.77 [0.98-3.19] | 0.057 | 2.47 [1.11-5.76] | 0.029 | 1.55 [0.9-2.65] | 0.11 |
| 16p13.11 | ADHD | 2.26 [1.37-3.73] | 0.0014 | 2.01 [1.43-2.81] | 5.0E-05 | 1.99 [1.24-3.2] | 0.0043 | 1.98 [1.44-2.74] | 2.90E-05 |
| 16p12.1 | ADHD | 1.86 [1.22-2.84] | 0.0039 | 0.56 [0.24-1.33] | 0.19 | 1.85 [1.25-2.74] | 0.0022 | 0.56 [0.22-1.28] | 0.19 |
| 16p11.2d | ADHD | 1.26 [0.55-2.88] | 0.58 | 1.57 [0.78-3.18] | 0.21 | 1.03 [0.45-2.27] | 0.95 | 1.69 [0.86-3.31] | 0.12 |
| 16p11.2 | ADHD | 0.77 [0.36-1.61] | 0.48 | 2.63 [1.75-3.95] | 3.3E-06 | 0.93 [0.42-1.92] | 0.84 | 3.13 [2.13-4.65] | 9.80E-09 |
| 17p12 | ADHD | 0.45 [0.18-1.15] | 0.095 | 0.69 [0.21-2.23] | 0.53 | 0.41 [0.15-0.98] | 0.061 | 0.71 [0.19-2.21] | 0.57 |
| 17q12 | ADHD | 4.24 [1.29-13.9] | 0.017 | 4.19 [1.98-8.87] | 0.0002 | 2.12 [0.72-6.56] | 0.17 | 3.66 [1.85-7.65] | 0.0003 |
| 22q11.2 | ADHD | 1.17 [0.49-2.8] | 0.72 | 2.61 [1.67-4.09] | 2.6E-05 | 1.48 [0.53-4.13] | 0.45 | 2.58 [1.69-3.99] | 1.40E-05 |
| 22q11.2b | ADHD | 1.81 [0.53-6.18] | 0.35 | 1.63 [0.55-4.83] | 0.38 | 1.97 [0.6-6.49] | 0.25 | 2.01 [0.7-5.92] | 0.19 |
| TAR | Any affective disorder | 0.9 [0.26-3.06] | 0.86 | 1.37 [0.87-2.18] | 0.18 | 1.01 [0.32-2.87] | 0.99 | 1.4 [0.91-2.18] | 0.13 |
| 1q21.1 | Any affective disorder | 2.13 [0.89-5.09] | 0.087 | 1.45 [0.76-2.77] | 0.25 | 1.83 [0.85-4.1] | 0.13 | 1.19 [0.65-2.15] | 0.57 |
| 2q11.2 | Any affective disorder | 2.06 [0.57-7.43] | 0.27 | na. | na. | 3.03 [0.62-21.99] | 0.20 | na. | na. |
| 2q13 | Any affective disorder | 1.29 [0.56-2.99] | 0.56 | 0.73 [0.19-2.82] | 0.65 | 0.85 [0.39-1.82] | 0.69 | 0.67 [0.18-2.26] | 0.52 |
| 2q21.1 | Any affective disorder | 1.51 [0.17-13.2] | 0.71 | 0.65 [0.17-2.53] | 0.53 | 0.88 [0.11-5.15] | 0.89 | 0.61 [0.15-2.12] | 0.45 |
| 10q11.23 | Any affective disorder | 0.72 [0.21-2.48] | 0.60 | 1.4 [0.3-6.57] | 0.67 | 0.57 [0.17-1.75] | 0.34 | 0.85 [0.16-3.98] | 0.83 |
| 13q12.12 | Any affective disorder | 0.36 [0.1-1.32] | 0.12 | 3.03 [0.66-13.9] | 0.15 | 0.44 [0.11-1.64] | 0.22 | 1.99 [0.54-7.61] | 0.30 |
| 15q11.2 | Any affective disorder | 1.15 [0.89-1.49] | 0.28 | 1.09 [0.84-1.4] | 0.51 | 1.11 [0.88-1.41] | 0.37 | 1.09 [0.86-1.38] | 0.49 |
| PWAS | Any affective disorder | na. | na. | 1.7 [0.29-10.1] | 0.56 | na. | na. | 0.68 [0.12-3.49] | 0.64 |
| 15q13.3 | Any affective disorder | 2.01 [0.79-5.13] | 0.14 | 1.48 [0.82-2.66] | 0.19 | 1.67 [0.74-3.98] | 0.22 | 1.53 [0.89-2.66] | 0.13 |
| 16p13.11 | Any affective disorder | 1.14 [0.61-2.14] | 0.69 | 1.42 [0.97-2.06] | 0.069 | 1.21 [0.7-2.08] | 0.50 | 1.56 [1.1-2.21] | 0.012 |
| 16p12.1 | Any affective disorder | 0.99 [0.6-1.62] | 0.96 | 0.87 [0.41-1.85] | 0.71 | 0.85 [0.54-1.35] | 0.49 | 0.96 [0.46-1.97] | 0.92 |
| 16p11.2d | Any affective disorder | 0.97 [0.37-2.54] | 0.95 | 1.2 [0.58-2.45] | 0.63 | 0.91 [0.38-2.15] | 0.84 | 1.23 [0.63-2.41] | 0.55 |
| 16p11.2 | Any affective disorder | 0.45 [0.2-1.03] | 0.057 | 0.99 [0.6-1.65] | 0.98 | 0.43 [0.17-0.98] | 0.054 | 1.1 [0.68-1.78] | 0.71 |

| | | | | | | | | | |
|---|---|---|---|---|---|---|---|---|---|
| 17p12 | Any affective disorder | 0.79 [0.35-1.77] | 0.57 | 0.52 [0.17-1.59] | 0.25 | 0.74 [0.34-1.6] | 0.44 | 0.64 [0.2-1.89] | 0.43 |
| 17q12 | Any affective disorder | 0.63 [0.08-5.11] | 0.68 | 1.56 [0.57-4.26] | 0.38 | 0.64 [0.09-3.14] | 0.62 | 1.98 [0.86-4.76] | 0.11 |
| 22q11.2 | Any affective disorder | 0.95 [0.28-3.24] | 0.93 | 0.98 [0.57-1.7] | 0.95 | 0.83 [0.26-2.51] | 0.74 | 1.06 [0.62-1.8] | 0.82 |
| 22q11.2b | Any affective disorder | 1.03 [0.24-4.49] | 0.97 | 1.84 [0.66-5.16] | 0.25 | 1.45 [0.39-5.95] | 0.59 | 1.67 [0.6-4.85] | 0.33 |
| TAR | Any iPSYCH disorder | 0.68 [0.29-1.6] | 0.38 | 1.48 [1.02-2.14] | 0.037 | 0.54 [0.24-1.21] | 0.13 | 1.61 [1.15-2.29] | 0.0074 |
| 1q21.1 | Any iPSYCH disorder | 2.12 [1.08-4.14] | 0.028 | 2.54 [1.58-4.08] | 0.0001 | 2.36 [1.3-4.63] | 0.0076 | 2.32 [1.53-3.66] | 0.0002 |
| 2q11.2 | Any iPSYCH disorder | 2.76 [0.93-8.12] | 0.067 | 0.31 [0.05-1.9] | 0.21 | 3.99 [1.12-25.35] | 0.067 | 0.26 [0.04-1.34] | 0.12 |
| 2q13 | Any iPSYCH disorder | 1.28 [0.69-2.38] | 0.43 | 1.1 [0.43-2.82] | 0.85 | 0.98 [0.57-1.72] | 0.94 | 1.1 [0.48-2.7] | 0.83 |
| 2q21.1 | Any iPSYCH disorder | 2.34 [0.61-8.87] | 0.21 | 0.45 [0.14-1.45] | 0.18 | 1.87 [0.67-6.6] | 0.27 | 0.39 [0.13-1.14] | 0.086 |
| 10q11.23 | Any iPSYCH disorder | 0.71 [0.28-1.78] | 0.47 | 1.63 [0.44-6.05] | 0.46 | 0.64 [0.28-1.52] | 0.3 | 1.13 [0.36-4.2] | 0.84 |
| 13q12.12 | Any iPSYCH disorder | 0.71 [0.26-1.91] | 0.50 | 3.81 [1.25-11.6] | 0.019 | 1.15 [0.42-3.63] | 0.79 | 2.03 [0.8-6.16] | 0.16 |
| 15q11.2 | Any iPSYCH disorder | 1.3 [1.07-1.58] | 0.008 | 1.21 [1-1.47] | 0.053 | 1.35 [1.13-1.61] | 0.0012 | 1.13 [0.95-1.36] | 0.16 |
| PWAS | Any iPSYCH disorder | na. | na. | 11.3 [4.24-30.1] | 1.3E-06 | na. | na. | 6.42 [2.6-21.36] | 0.0004 |
| 15q13.3 | Any iPSYCH disorder | 2.7 [1.31-5.56] | 0.007 | 1.53 [0.94-2.5] | 0.091 | 2.69 [1.43-5.62] | 0.0042 | 1.38 [0.9-2.16] | 0.15 |
| 16p13.11 | Any iPSYCH disorder | 1.75 [1.11-2.76] | 0.017 | 1.7 [1.27-2.27] | 0.0003 | 1.63 [1.1-2.48] | 0.019 | 1.73 [1.33-2.3] | 7.70E-05 |
| 16p12.1 | Any iPSYCH disorder | 1.26 [0.86-1.84] | 0.24 | 0.81 [0.45-1.44] | 0.47 | 1.19 [0.86-1.69] | 0.30 | 0.86 [0.48-1.59] | 0.62 |
| 16p11.2d | Any iPSYCH disorder | 1.23 [0.62-2.44] | 0.56 | 1.32 [0.73-2.4] | 0.36 | 1.13 [0.62-2.14] | 0.71 | 1.39 [0.82-2.45] | 0.23 |
| 17p12 | Any iPSYCH disorder | 0.61 [0.33-1.15] | 0.13 | 0.64 [0.27-1.51] | 0.31 | 0.64 [0.35-1.16] | 0.13 | 0.73 [0.33-1.71] | 0.46 |
| 22q11.2 | Any iPSYCH disorder | 1.85 [0.83-4.14] | 0.13 | 1.85 [1.23-2.8] | 0.0035 | 2.04 [0.94-5.07] | 0.093 | 2 [1.37-3] | 0.0005 |
| 22q11.2b | Any iPSYCH disorder | 0.64 [0.2-2.03] | 0.37 | 1.58 [0.64-3.89] | 0.32 | 1.98 [0.78-6.01] | 0.18 | 1.53 [0.68-3.88] | 0.33 |
| TAR | ASD | 0.64 [0.2-2.03] | 0.44 | 1.49 [0.95-2.34] | 0.084 | 0.5 [0.13-1.52] | 0.25 | 1.61 [1.02-2.52] | 0.04 |
| 1q21.1 | ASD | 2.37 [1.16-4.86] | 0.018 | 4.03 [2.44-6.64] | 4.6E-08 | 2.77 [1.3-6.12] | 0.0092 | 4.1 [2.52-6.84] | 2.80E-08 |
| 2q11.2 | ASD | 2.44 [0.54-11] | 0.25 | na. | na. | 1.38 [0.25-6.94] | 0.70 | na. | na. |
| 2q13 | ASD | 0.92 [0.4-2.08] | 0.84 | 0.97 [0.27-3.45] | 0.97 | 0.8 [0.33-1.76] | 0.59 | 0.96 [0.24-3.32] | 0.95 |
| 2q21.1 | ASD | 2.49 [0.64-9.8] | 0.19 | 0.61 [0.12-3.14] | 0.55 | 2.16 [0.6-8.59] | 0.24 | 0.56 [0.08-2.38] | 0.47 |
| 10q11.23 | ASD | 0.74 [0.22-2.51] | 0.63 | 1.81 [0.35-9.35] | 0.48 | 0.81 [0.21-2.57] | 0.73 | 1.82 [0.34-8.74] | 0.45 |
| 13q12.12 | ASD | 0.77 [0.15-3.98] | 0.75 | 5 [1.47-16.99] | 0.010 | 0.86 [0.11-4.39] | 0.87 | 2.66 [0.8-9.42] | 0.11 |
| 15q11.2 | ASD | 1.45 [1.15-1.84] | 0.0018 | 1.07 [0.82-1.38] | 0.63 | 1.52 [1.2-1.92] | 0.0005 | 0.94 [0.72-1.21] | 0.62 |
| PWAS | ASD | na. | na. | 20.8 [7.86-55] | 9.6E-10 | na. | na. | 19.25 [7.52-65.35] | 3.70E-08 |
| 15q13.3 | ASD | 3.74 [1.66-8.42] | 0.0014 | 1.59 [0.85-2.98] | 0.15 | 4.64 [2.15-10.66] | 0.0001 | 1.51 [0.81-2.76] | 0.18 |
| 16p13.11 | ASD | 1.92 [1.12-3.31] | 0.018 | 1.84 [1.30-2.61] | 0.0006 | 1.65 [0.97-2.81] | 0.064 | 1.76 [1.24-2.49] | 0.0015 |
| 16p12.1 | ASD | 1.11 [0.68-1.83] | 0.68 | 1.00 [0.46-2.19] | 1 | 1.1 [0.67-1.79] | 0.69 | 0.94 [0.41-2.02] | 0.88 |
| 16p11.2d | ASD | 0.94 [0.37-2.39] | 0.89 | 1.26 [0.57-2.79] | 0.57 | 0.84 [0.31-2.11] | 0.73 | 1.28 [0.56-2.8] | 0.55 |
| 16p11.2 | ASD | 3.07 [1.75-5.36] | 8.4E-05 | 2.04 [1.33-3.15] | 0.0012 | 3.9 [2.22-7.03] | 3.20E-06 | 2.52 [1.61-3.96] | 5.30E-05 |
| 17p12 | ASD | 0.81 [0.36-1.82] | 0.61 | 0.64 [0.17-2.41] | 0.51 | 0.74 [0.31-1.65] | 0.48 | 0.59 [0.13-2.12] | 0.45 |
| 17q12 | ASD | 7.79 [2.71-22.4] | 1.40E-04 | 3.12 [1.37-7.08] | 0.0065 | 3.86 [1.52-10.6] | 0.0056 | 3.04 [1.35-7.04] | 0.0079 |
| 22q11.2 | ASD | 1.5 [0.59-3.84] | 0.39 | 2.63 [1.66-4.16] | 3.4E-05 | 1.53 [0.57-4.09] | 0.39 | 2.65 [1.67-4.24] | 4.30E-05 |
| 22q11.2b | ASD | 2.34 [0.67-8.13] | 0.18 | 0.92 [0.25-3.30] | 0.89 | 2.55 [0.74-8.8] | 0.13 | 1.12 [0.29-3.85] | 0.86 |

| | | | | | | | | | |
|---|---|---|---|---|---|---|---|---|---|
| 1q21.1 | Bipolar disorder | 3.52 [0.88-14.1] | 0.076 | 1.63 [0.37-7.32] | 0.52 | 3.48 [0.75-11.85] | 0.067 | 1.3 [0.21-4.52] | 0.72 |
| 15q11.2 | Bipolar disorder | 0.63 [0.3-1.31] | 0.22 | 1.26 [0.72-2.20] | 0.41 | 0.65 [0.29-1.26] | 0.25 | 1.23 [0.68-2.04] | 0.46 |
| 15q13.3 | Bipolar disorder | na. | na. | 0.85 [0.19-3.79] | 0.84 | na. | na. | 0.86 [0.14-2.99] | 0.84 |
| 16p13.11 | Bipolar disorder | 2.83 [1.11-7.23] | 0.029 | 0.8 [0.28-2.27] | 0.67 | 2.86 [1.1-6.58] | 0.02 | 0.8 [0.24-1.98] | 0.67 |
| 16p12.1 | Bipolar disorder | 1.62 [0.65-4.03] | 0.30 | na. | na. | 1.65 [0.61-3.71] | 0.27 | na. | na. |
| 16p11.2d | Bipolar disorder | na. | na. | 1.89 [0.51-6.93] | 0.34 | na. | na. | 1.92 [0.44-5.91] | 0.31 |
| 16p11.2 | Bipolar disorder | na. | na. | 0.53 [0.13-2.28] | 0.40 | na. | na. | 0.6 [0.1-2.02] | 0.49 |
| 17q12 | Bipolar disorder | na. | na. | 2.15 [0.39-11.8] | 0.38 | na. | na. | 2.29 [0.33-9.86] | 0.31 |
| TAR | Epilepsy | na. | na. | 1.49 [0.65-3.42] | 0.34 | na. | na. | 0.99 [0.39-2.06] | 0.99 |
| 1q21.1 | Epilepsy | 1.63 [0.41-6.47] | 0.49 | 1.79 [0.66-4.88] | 0.25 | 1.18 [0.29-3.2] | 0.79 | 0.68 [0.21-1.63] | 0.45 |
| 2q11.2 | Epilepsy | 9.52 [2.42-37.4] | 0.0013 | na. | na. | 3.39 [0.53-12.3] | 0.11 | na. | na. |
| 2q13 | Epilepsy | 0.64 [0.09-4.69] | 0.66 | na. | na. | 0.51 [0.03-2.33] | 0.50 | na. | na. |
| 2q21.1 | Epilepsy | 4.17 [0.48-36.1] | 0.2 | na. | na. | 3.49 [0.54-12.73] | 0.10 | na. | na. |
| 10q11.23 | Epilepsy | na. | na. | 3.15 [0.36-27.8] | 0.30 | na. | na. | 2.39 [0.13-12.34] | 0.40 |
| 13q12.12 | Epilepsy | 3.2 [0.65-15.8] | 0.15 | 3.24 [0.59-17.7] | 0.17 | 3.65 [0.57-13.14] | 0.088 | 2.49 [0.39-8.66] | 0.22 |
| 15q11.2 | Epilepsy | 1.90 [1.29-2.79] | 0.001 | 0.91 [0.53-1.55] | 0.72 | 1.69 [1.17-2.35] | 0.0031 | 0.73 [0.41-1.19] | 0.24 |
| PWAS | Epilepsy | na. | na. | 15.1 [7.38-30.9] | 1.1E-13 | na. | na. | 3.73 [1.67-7.5] | 0.0005 |
| 15q13.3 | Epilepsy | 7.33 [3.32-16.2] | 8.1E-07 | 1.70 [0.62-4.62] | 0.30 | 3.45 [1.5-6.91] | 0.0013 | 1.62 [0.63-3.44] | 0.26 |
| 16p13.11 | Epilepsy | 10.0 [6.60-15.2] | 3.2E-27 | 0.50 [0.16-1.56] | 0.23 | 9.3 [6.05-13.92] | 6.10E-26 | 0.35 [0.11-0.84] | 0.04 |
| 16p12.1 | Epilepsy | 1.64 [0.76-3.56] | 0.21 | 0.63 [0.09-4.65] | 0.65 | 1.39 [0.62-2.67] | 0.37 | 0.68 [0.04-3.16] | 0.70 |
| 16p11.2d | Epilepsy | 5.26 [2.17-12.8] | 2.40E-04 | 1.20 [0.28-5.07] | 0.81 | 4.13 [1.56-9.17] | 0.0014 | 0.83 [0.14-2.68] | 0.80 |
| 16p11.2 | Epilepsy | 5.47 [2.59-11.6] | 8.7E-06 | 2.35 [1.20-4.62] | 0.013 | 4.06 [2.07-7.31] | 1.10E-05 | 1.4 [0.69-2.54] | 0.30 |
| 17p12 | Epilepsy | na. | na. | 3.35 [1.00-11.2] | 0.050 | na. | na. | 4.34 [1.01-12.88] | 0.019 |
| 17q12 | Epilepsy | na. | na. | 2.01 [0.48-8.45] | 0.34 | na. | na. | 0.84 [0.14-2.71] | 0.81 |
| 22q11.2 | Epilepsy | 11.4 [5.36-24.4] | 2.8E-10 | 1.97 [0.85-4.60] | 0.12 | 7.43 [3.11-15.92] | 1.10E-06 | 1.18 [0.5-2.35] | 0.67 |
| 22q11.2b | Epilepsy | 6.38 [2.03-20.1] | 0.0015 | 1.48 [0.20-11.2] | 0.70 | 4.01 [0.93-12] | 0.028 | 2.04 [0.33-6.9] | 0.34 |
| TAR | Intellectual disability | 1.18 [0.26-5.29] | 0.83 | 1.18 [0.55-2.51] | 0.67 | 1.45 [0.21-5.8] | 0.64 | 0.76 [0.33-1.49] | 0.46 |
| 1q21.1 | Intellectual disability | 5.38 [2.40-12.09] | 4.6E-05 | 3.14 [1.59-6.20] | 9.90E-04 | 2.99 [1.41-5.83] | 0.0023 | 1.1 [0.57-1.96] | 0.75 |
| 2q11.2 | Intellectual disability | 13.43 [2.97-60.7] | 7.40E-04 | na. | na. | 3.79 [0.82-12.83] | 0.05 | na. | na. |
| 2q13 | Intellectual disability | 1.54 [0.52-4.56] | 0.44 | 0.83 [0.10-6.68] | 0.86 | 1.24 [0.36-3.26] | 0.69 | 0.85 [0.05-4.14] | 0.87 |
| 2q21.1 | Intellectual disability | 4.67 [1.27-17.2] | 0.021 | 2.05 [0.41-10.1] | 0.38 | 2.76 [0.61-9.03] | 0.13 | 4.58 [0.62-20.17] | 0.075 |
| 10q11.23 | Intellectual disability | 0.67 [0.08-5.29] | 0.70 | 1.97 [0.22-17.72] | 0.54 | 1.08 [0.06-5.41] | 0.94 | 1.95 [0.1-10.57] | 0.53 |
| 13q12.12 | Intellectual disability | 2.4 [0.48-12.1] | 0.29 | 2.09 [0.25-17.3] | 0.49 | 2.32 [0.33-9.67] | 0.31 | 0.69 [0.04-3.52] | 0.73 |
| 15q11.2 | Intellectual disability | 1.46 [1.02-2.07] | 0.038 | 1.25 [0.86-1.83] | 0.24 | 1.14 [0.8-1.58] | 0.45 | 1.09 [0.74-1.55] | 0.66 |
| PWAS | Intellectual disability | na. | na. | 46.5 [20.1-108] | 3.5E-19 | na. | na. | 6.85 [3.7-12.56] | 5.40E-10 |
| 15q13.3 | Intellectual disability | 10.4 [5.03-21.5] | 2.7E-10 | 2.28 [1.04-5] | 0.041 | 6.9 [3.72-12.32] | 2.00E-10 | 1.47 [0.63-2.98] | 0.33 |
| 16p13.11 | Intellectual disability | 6.12 [3.63-10.33] | 1.10E-11 | 2.04 [1.24-3.35] | 0.0051 | 4.24 [2.58-6.72] | 3.20E-09 | 1.15 [0.69-1.8] | 0.56 |
| 16p12.1 | Intellectual disability | 1.49 [0.75-2.96] | 0.25 | 1.24 [0.36-4.26] | 0.73 | 1.11 [0.53-2.07] | 0.75 | 1.52 [0.36-4.33] | 0.50 |

| | | | | | | | | | |
|---|---|---|---|---|---|---|---|---|---|
| 16p11.2d | Intellectual disability | 2.76 [1.07-7.11] | 0.036 | 1.86 [0.69-5.01] | 0.22 | 2.94 [1.08-6.73] | 0.019 | 1.52 [0.52-3.57] | 0.38 |
| 16p11.2 | Intellectual disability | 5.14 [2.67-9.91] | 1.0E-06 | 4.43 [2.76-7.09] | 5.9E-10 | 3.69 [2.02-6.37] | 7.70E-06 | 2.65 [1.7-4.01] | 8.30E-06 |
| 17p12 | Intellectual disability | 0.33 [0.04-2.49] | 0.28 | 2.80 [0.88-8.86] | 0.080 | 0.43 [0.02-2.08] | 0.41 | 4.66 [1.28-13.28] | 0.0082 |
| 17q12 | Intellectual disability | 16.1 [5.90-43.8] | 5.60E-08 | 2.96 [1.06-8.25] | 0.038 | 5.66 [2.34-12.77] | 5.10E-05 | 1.23 [0.42-2.87] | 0.67 |
| 22q11.2 | Intellectual disability | 18.5 [7.98-42.7] | 9.6E-12 | 3.80 [2.15-6.71] | 4.4E-06 | 31.79 [15.7-65.8] | 1.60E-21 | 1.79 [1.03-2.92] | 0.028 |
| 22q11.2b | Intellectual disability | 1.66 [0.20-13.8] | 0.64 | na. | na. | 0.71 [0.04-3.57] | 0.75 | na. | na. |
| TAR | Major depressive disorder | 0.96 [0.28-3.28] | 0.95 | 1.45 [0.92-2.31] | 0.11 | 1.07 [0.35-3.06] | 0.90 | 1.48 [0.96-2.3] | 0.08 |
| 1q21.1 | Major depressive disorder | 2.08 [0.86-5.04] | 0.10 | 1.43 [0.74-2.75] | 0.28 | 1.7 [0.77-3.87] | 0.19 | 1.23 [0.67-2.24] | 0.51 |
| 2q11.2 | Major depressive disorder | 2.18 [0.61-7.86] | 0.23 | na. | na. | 3.2 [0.66-23.16] | 0.18 | na. | na. |
| 2q13 | Major depressive disorder | 1.39 [0.6-3.22] | 0.44 | 0.79 [0.2-3.04] | 0.73 | 0.93 [0.42-1.98] | 0.86 | 0.71 [0.19-2.4] | 0.59 |
| 2q21.1 | Major depressive disorder | na. | na. | 0.52 [0.12-2.29] | 0.39 | na. | na. | 0.51 [0.11-1.91] | 0.34 |
| 10q11.23 | Major depressive disorder | 0.77 [0.22-2.68] | 0.69 | 1 [0.18-5.66] | 1.00 | 0.62 [0.18-1.87] | 0.41 | 0.63 [0.08-3.36] | 0.60 |
| 13q12.12 | Major depressive disorder | 0.39 [0.11-1.42] | 0.15 | 2.67 [0.56-12.8] | 0.22 | 0.48 [0.12-1.77] | 0.27 | 1.7 [0.42-6.84] | 0.44 |
| 15q11.2 | Major depressive disorder | 1.17 [0.91-1.52] | 0.22 | 1.07 [0.82-1.38] | 0.62 | 1.14 [0.9-1.44] | 0.29 | 1.05 [0.83-1.34] | 0.68 |
| PWAS | Major depressive disorder | na. | na. | 1.84 [0.31-10.9] | 0.50 | na. | na. | 0.73 [0.13-3.71] | 0.70 |
| 15q13.3 | Major depressive disorder | 2.16 [0.84-5.5] | 0.11 | 1.54 [0.86-2.78] | 0.15 | 1.79 [0.79-4.25] | 0.17 | 1.57 [0.91-2.74] | 0.11 |
| 16p13.11 | Major depressive disorder | 1.06 [0.56-2.03] | 0.85 | 1.38 [0.94-2.01] | 0.10 | 1.14 [0.65-1.99] | 0.65 | 1.55 [1.09-2.22] | 0.016 |
| 16p12.1 | Major depressive disorder | 0.91 [0.54-1.52] | 0.71 | 0.86 [0.4-1.87] | 0.71 | 0.77 [0.48-1.24] | 0.29 | 0.96 [0.45-1.99] | 0.91 |
| 16p11.2d | Major depressive disorder | 1.04 [0.4-2.73] | 0.94 | 1.17 [0.56-2.42] | 0.67 | 1.01 [0.42-2.39] | 0.98 | 1.22 [0.62-2.42] | 0.57 |
| 16p11.2 | Major depressive disorder | 0.42 [0.18-1] | 0.049 | 1.04 [0.63-1.74] | 0.87 | 0.4 [0.15-0.94] | 0.044 | 1.16 [0.71-1.87] | 0.56 |
| 17p12 | Major depressive disorder | 0.85 [0.38-1.91] | 0.70 | 0.55 [0.18-1.71] | 0.30 | 0.82 [0.37-1.77] | 0.60 | 0.68 [0.22-2.02] | 0.50 |
| 17q12 | Major depressive disorder | na. | na. | 1.47 [0.53-4.07] | 0.46 | na. | na. | 1.78 [0.75-4.37] | 0.19 |
| 22q11.2 | Major depressive disorder | 1.03 [0.3-3.52] | 0.96 | 1.02 [0.59-1.77] | 0.94 | 0.89 [0.27-2.68] | 0.83 | 1.11 [0.65-1.89] | 0.70 |
| 22q11.2b | Major depressive disorder | 1.11 [0.25-4.81] | 0.89 | 1.32 [0.43-4.08] | 0.63 | 1.54 [0.41-6.33] | 0.52 | 1.09 [0.33-3.44] | 0.88 |
| TAR | Schizophrenia | na. | na. | 1.5 [0.77-2.93] | 0.23 | na. | na. | 1.64 [0.81-3.13] | 0.15 |
| 1q21.1 | Schizophrenia | 1.75 [0.52-5.84] | 0.36 | 2.36 [0.97-5.71] | 0.058 | 1.65 [0.43-5.13] | 0.42 | 2.52 [1.02-5.58] | 0.031 |
| 2q11.2 | Schizophrenia | 7.77 [1.25-48.2] | 0.028 | na. | na. | 7.75 [1.32-45.64] | 0.018 | na. | na. |
| 2q13 | Schizophrenia | 1.39 [0.37-5.18] | 0.63 | 1.63 [0.31-8.59] | 0.57 | 1.27 [0.28-3.99] | 0.72 | 2.07 [0.3-9.48] | 0.39 |
| 10q11.23 | Schizophrenia | 1.3 [0.25-6.66] | 0.76 | na. | na. | 1.18 [0.17-4.96] | 0.84 | na. | na. |
| 13q12.12 | Schizophrenia | na. | na. | 12.7 [2.17-74.1] | 0.0048 | na. | na. | 7.08 [1.33-31.34] | 0.012 |
| 15q11.2 | Schizophrenia | 1.29 [0.88-1.88] | 0.19 | 1.25 [0.84-1.87] | 0.27 | 1.36 [0.92-1.95] | 0.11 | 1.2 [0.8-1.76] | 0.36 |
| PWAS | Schizophrenia | na. | na. | 6.88 [0.95-49.8] | 0.056 | na. | na. | 7.64 [1.41-37.46] | 0.012 |
| 15q13.3 | Schizophrenia | 3.28 [1.02-10.6] | 0.047 | 1.3 [0.5-3.37] | 0.59 | 3.33 [1.01-9.69] | 0.033 | 1.27 [0.45-3.06] | 0.62 |
| 16p13.11 | Schizophrenia | 1.77 [0.78-4.01] | 0.17 | 2.05 [1.22-3.45] | 0.0069 | 1.85 [0.77-4.02] | 0.14 | 1.95 [1.15-3.22] | 0.011 |
| 16p12.1 | Schizophrenia | 1.34 [0.64-2.8] | 0.44 | 0.65 [0.15-2.87] | 0.56 | 1.3 [0.59-2.58] | 0.49 | 0.67 [0.1-2.37] | 0.59 |
| 16p11.2d | Schizophrenia | 2.42 [0.78-7.52] | 0.13 | 2.38 [0.95-5.91] | 0.063 | 2.3 [0.7-6.54] | 0.14 | 2.66 [1.05-6.19] | 0.028 |
| 16p11.2 | Schizophrenia | 0.57 [0.13-2.5] | 0.45 | 1.22 [0.59-2.52] | 0.58 | 0.61 [0.1-2.14] | 0.51 | 1.36 [0.63-2.71] | 0.40 |
| 17p12 | Schizophrenia | na. | na. | 2.38 [0.69-8.2] | 0.17 | na. | na. | 3.21 [0.84-10.25] | 0.06 |

| | | | | | | | | | |
|---|---|---|---|---|---|---|---|---|---|
| 17q12 | Schizophrenia | 5.38 [0.61-47.2] | 0.13 | 3.25 [0.98-10.7] | 0.054 | 3.73 [0.48-20.44] | 0.15 | 2.8 [0.82-8.69] | 0.08 |
| 22q11.2 | Schizophrenia | 3.18 [0.92-10.9] | 0.067 | 2.02 [0.99-4.12] | 0.053 | 2.35 [0.67-7.46] | 0.16 | 2.19 [1.05-4.3] | 0.028 |
| 22q11.2b | Schizophrenia | na. | na. | 2.56 [0.59-11] | 0.21 | na. | na. | 2.82 [0.59-10.75] | 0.15 |
| TAR | Schizophrenia spectrum disorder | 1.63 [0.45-5.83] | 0.45 | 1.02 [0.56-1.86] | 0.95 | 1.51 [0.4-4.76] | 0.50 | 1.08 [0.57-1.93] | 0.81 |
| 1q21.1 | Schizophrenia spectrum disorder | 1.78 [0.68-4.69] | 0.24 | 2.7 [1.37-5.33] | 0.0042 | 1.64 [0.61-4.15] | 0.30 | 2.47 [1.3-4.61] | 0.0049 |
| 2q11.2 | Schizophrenia spectrum disorder | 4.75 [1.07-21.1] | 0.041 | na. | na. | 8.3 [1.46-64.14] | 0.021 | na. | na. |
| 2q13 | Schizophrenia spectrum disorder | 1.58 [0.59-4.19] | 0.36 | 1.23 [0.29-5.18] | 0.78 | 1.15 [0.42-2.85] | 0.77 | 1.5 [0.32-5.47] | 0.56 |
| 2q21.1 | Schizophrenia spectrum disorder | 2.5 [0.28-22.8] | 0.42 | na. | na. | 2.21 [0.27-13.19] | 0.40 | na. | na. |
| 10q11.23 | Schizophrenia spectrum disorder | 0.66 [0.13-3.3] | 0.61 | 3.22 [0.6-17.1] | 0.17 | 0.62 [0.09-2.6] | 0.56 | 1.81 [0.32-9.09] | 0.47 |
| 15q11.2 | Schizophrenia spectrum disorder | 0.99 [0.71-1.36] | 0.93 | 1.32 [0.97-1.78] | 0.077 | 1.03 [0.74-1.41] | 0.86 | 1.25 [0.92-1.67] | 0.14 |
| PWAS | Schizophrenia spectrum disorder | na. | na. | 9.19 [1.72-49.2] | 0.010 | na. | na. | 8.03 [2.48-28.41] | 0.0006 |
| 15q13.3 | Schizophrenia spectrum disorder | 3.49 [1.36-8.95] | 0.0093 | 1.44 [0.71-2.95] | 0.31 | 2.95 [1.19-7.39] | 0.018 | 1.52 [0.72-3.07] | 0.25 |
| 16p13.11 | Schizophrenia spectrum disorder | 2.29 [1.22-4.3] | 0.010 | 1.98 [1.31-3] | 0.0012 | 2.2 [1.19-4.03] | 0.011 | 1.99 [1.31-3] | 0.0011 |
| 16p12.1 | Schizophrenia spectrum disorder | 1.21 [0.66-2.2] | 0.54 | 0.48 [0.14-1.68] | 0.25 | 1.2 [0.65-2.12] | 0.55 | 0.45 [0.1-1.36] | 0.21 |
| 16p11.2d | Schizophrenia spectrum disorder | 2.41 [0.96-6.08] | 0.062 | 1.81 [0.81-4.04] | 0.15 | 2.03 [0.82-4.89] | 0.12 | 2.07 [0.94-4.42] | 0.062 |
| 16p11.2 | Schizophrenia spectrum disorder | 1.26 [0.55-2.88] | 0.58 | 1.5 [0.89-2.54] | 0.13 | 1.28 [0.54-2.79] | 0.56 | 1.86 [1.07-3.16] | 0.024 |
| 17p12 | Schizophrenia spectrum disorder | 0.46 [0.13-1.65] | 0.24 | 1.17 [0.34-3.99] | 0.80 | 0.48 [0.11-1.48] | 0.25 | 1.54 [0.41-4.85] | 0.48 |
| 17q12 | Schizophrenia spectrum disorder | 4.84 [0.81-28.9] | 0.084 | 3.19 [1.17-8.68] | 0.023 | 3.56 [0.8-15.12] | 0.083 | 2.92 [1.13-7.59] | 0.025 |
| 22q11.2 | Schizophrenia spectrum disorder | 3.28 [1.15-9.3] | 0.026 | 1.34 [0.7-2.54] | 0.37 | 2.41 [0.91-6.51] | 0.077 | 1.46 [0.75-2.71] | 0.25 |
| 22q11.2b | Schizophrenia spectrum disorder | 2.6 [0.65-10.4] | 0.18 | 2.51 [0.77-8.16] | 0.12 | 2.64 [0.7-9.88] | 0.14 | 2.75 [0.85-8.67] | 0.08 |
